# Effect of maternal and infant selenium status on child growth in a birth cohort from Dhaka, Bangladesh

**DOI:** 10.1101/2022.11.01.22281815

**Authors:** Rukshan Mehta, Christine Krupa, Tahmeed Ahmed, Davidson H. Hamer, Abdullah Al Mahmud

**Affiliations:** Centre for Global Child Health, The Hospital for Sick Children, Toronto, Canada; Department of Nutritional Sciences, Faculty of Medicine, University of Toronto, Toronto, Canada; Nutrition and Clinical Services Division, International Centre for Diarrhoeal Disease Research, Bangladesh (icddr,b), Dhaka, Bangladesh; Department of Global Health, Boston University School of Public Health, Boston, MA, USA; Section of Infectious Diseases, Boston University School of Medicine, Boston, MA, USA; Gerald J. and Dorothy R. Friedman School of Nutrition Science and Policy, Tufts University, Boston, MA, USA

## Abstract

Deficiency of selenium, an essential trace element, has been implicated in adverse birth outcomes and the growth of infants and young children.

We used data from a randomized controlled trial to examine associations between selenium biomarkers in whole blood (WBSe), serum and selenoprotein P (SEPP1) in maternal delivery and venous cord (VC) blood, and birth weight, and adverse birth outcomes. Furthermore, we examined associations between selenium biomarkers and infant growth outcomes (age adjusted length, weight, head circumference and weight-for-length z-scores) at birth, one, and two years of age using linear regression.

WB and serum selenium in delivery and VC specimens were negatively associated with birth weight (adjusted β, 95% CI: WBSe delivery: -26.6 (−44.3, -8.9); WBSe VC: -19.6 (−33.0, -6.1)); however, delivery SEPP1 levels (adjusted β: -37.5 (−73.0, -2.0)) and VC blood (adjusted β: 82.3 (30.0, 134.7)) showed inconsistent associations across biomarkers. We found small to moderate associations between infant growth and WBSe VC (LAZ β, 95% CI, at birth: -0.05 (−0.1, -0.01)); 12-months (β: -0.05 (−0.08, -0.007)). WAZ also showed weak negative associations with delivery WBSe (at birth: -0.07 (−0.1, -0.02); 12-months: -0.05 (−0.1, -0.005)) and in WBSe VC (β at birth: -0.05 (−0.08, -0.02); 12-months: -0.05 (−0.09, -0.004)).

Mechanisms connected to redox biology and its antioxidant effects have been causally associated with selenium’s protective properties. Given the fine balance between nutritional and toxic properties of selenium, it is possible that WB and serum selenium may negatively impact growth outcomes, both in utero and postpartum.

## Introduction

Linear growth, an important indicator of a child’s present and future health, is influenced by a complex interaction of environmental factors including intrauterine growth restriction, household socioeconomic status, parental education levels, inadequate maternal and child nutrition, and frequent infections.^1-2^ Global estimates (most recently from 2019) suggest that over 149 million children under the age of 5 years are stunted.^3^ According to the 2017-2018 Bangladesh Demographic and Health Survey (DHS), indicators of malnutrition among children under 5 years of age declined between 2007-2018, with the most recent prevalence estimates for stunting in children at 31%, 22% for underweight, and 8% for wasting.^4^

Selenium (Se) is an essential trace element that is known for its role in redox biology.^5^ Due to adaptive changes in selenium metabolism that occur during pregnancy, requirements are higher due to its role in fetal growth, which manifests as lower maternal concentrations of blood selenium.^6^ Concentrations of this trace element are known to fall during pregnancy, with lowest levels observed at delivery.^7-8^ Levels are lower because of increased fetal demands and alterations in intestinal absorption or renal handling.^7-8^ Reduced selenium concentrations can have an adverse impact on antioxidant selenoproteins, which compromises the protection against placental oxidative stress during pregnancy, negatively impacting fetal growth.^7^ Selenium is known to play a protective role in many birth outcomes.^9^ A recent meta-analysis found that umbilical cord blood selenium concentrations (r: 0.08, 95% CI: 0.01-0.16) are correlated with birth weight; although similar results were not observed for associations between maternal blood selenium and birthweight.^6^ Other studies have demonstrated significant relationships between low birth weight (LBW) and lower selenium concentrations.^6^ Mechanisms for the associations between birth weight and selenium remain unclear; however its antioxidant properties play an important role in placental development and oxidative balance.^6^ Lower selenium levels are likely associated with reduced function of glutathione peroxidase (GPx); an antioxidant enzyme involved in protecting the placenta and fetus from oxidative stress.^5,10^ In a similar manner, SEPP1 (selenoprotein P) is a selenoenzyme glycoprotein that protects against adverse pregnancy outcomes which can have an impact on fetal growth.^5,10^ Selenium levels may also be correlated with small-for-gestational age (SGA) status; mechanistically these associations may function via reductions in glucose levels which subsequently impact fetal weight.^11^ Lower placental GPx activity has been observed in women with SGA compared to controls.^12^ Due to the effects of lowered Se levels and decreased GPx activity during pregnancy, placental antioxidant defense may be reduced which in turn may impact fetal growth, resulting in adverse birth outcomes including growth restriction and SGA.^12^ A comprehensive schematic of placental to fetal transfer of various selenometabolites and Se metabolism in the human body is presented in **Supplementary Figure 1**.

In addition to the effects of selenium on birthweight and birth outcomes, this micronutrient also plays an essential role in bone growth and development. Selenium inadequacy has been implicated in growth faltering and changes in bone metabolism which can impact bone and cartilage development.^13-15^ Expression of selenoproteins by bone cells can contribute to protection against oxidative stress in the microenvironment of the bone.^15^ Selenium deficiency impacts bone formation and resorption, whereby the effects of selenium on bone are mediated through reduced antioxidative capacity, accumulation of reactive oxygen species, reduced osteoclast activation and osteoblast differentiation.^16^ Furthermore, selenium deficiency is known to impact cartilage and can reduce bone mineral content, as described in human and animal studies.^13-14,16^

Selenium metabolism and use in the bone is exclusively associated with selenoproteins. A total of nine selenoproteins have been identified in human fetal osteoblasts where they play a role in bone metabolism.^15^ SEPP1 is the primary transporter and storage selenoprotein from the liver to bones, where the Lrp8 receptor facilitates SEPP1-mediated Se uptake, particularly under conditions of Se deficiency. ^17-18^ Selenium therefore plays an important role in bone physiology and is tightly regulated by SEPP1 receptor, apolipoprotein receptor 2 (APOER2). Expression of this receptor is significantly influenced by SEPP1 levels in a negative feedback loop.^18^ Genetic ablation of SEPP1 has been shown to reduce serum Se concentrations by 25-fold, but only reduces bone Se levels by 2.5-fold, suggesting a protective effect.^18^

Studies have examined associations between selenium levels in mother-infant dyads and growth outcomes at birth and in the postnatal period.^6, 9, 19^ Given the mixed body of evidence on the association of selenium with birth weight and infant growth outcomes, we sought to understand whether selenium status measured in maternal and infant whole blood (WBSe), serum (serum Se) and a biomarker of functional status, selenoprotein P (SEPP1), at delivery and in venous cord blood are correlated with infant birth weight, growth outcomes (length-for-age, weight-for-age, weight-for-length and head circumference-for-age z-scores) at birth, in infancy, and up to 2 years of age. Our study will add to a growing body of evidence on the role selenium may play in in utero and postnatal infant growth and development.

## Methods

### Study Area & Subjects

We used observational selenium biomarker data collected as part of double-blinded, dose-varying, placebo-controlled, randomized trial of the effect of maternal vitamin D_3_ supplementation during pregnancy and lactation conducted in Dhaka, Bangladesh for this secondary analysis. Details of the parent study (NCT01924013) are described elsewhere (Roth et al. 2018).^20^ Briefly, a total of 1,300 women, 18 years of age and older, with uncomplicated singleton pregnancies were enrolled between 17-24 weeks of gestation and randomized into one of five treatment groups (treatment group A = 0 IU/week during pregnancy and postpartum, treatment group B = 4,200 IU/week during pregnancy and placebo postpartum, treatment group C = 16,800 IU/week during pregnancy and placebo postpartum, treatment group D = 28,000 IU/week during pregnancy and placebo postpartum, and treatment group E = 28,000 IU/week during pregnancy and postpartum).

Exclusion criteria included high risk pregnancies, women who had pre-existing medical conditions or those who were taking medications that could put them at risk of vitamin D sensitivity, altered vitamin D metabolism and hypercalcemia. Additional details are provided elsewhere.^20^

Inclusion of participant samples for this secondary analysis were based on availability of at least one measurement of selenium and/or selenoproteins at delivery in mothers or in venous cord blood of neonates. Infants for whom length and weight were not measured at birth or 12 months were excluded.

Selenium was measured as part of a panel of metals and minerals for which the primary study intent was to examine effects of vitamin D supplementation on heavy metal concentrations.^21^ Samples selected for Se quantification were not related to vitamin D treatment group and seasonal imbalances in specimen collection were observed for those selected to be a part of the sub-study.^21^ Additional sociodemographic characteristics of study participants were recorded using standardized data collection forms.

The parent study (MDIG: 1000057297) was conducted according to the guidelines laid down in the Declaration of Helsinki and all procedures involving human subjects and secondary specimen use sub-studies including the heavy metals protocol were approved by ethical review committees at the Hospital for Sick Children (Canada) and the International Center for Diarrheal Disease Research, Bangladesh. All women in the study provided written informed consent which included the use of biological specimens for future research purposes. This sub-study was approved by the ethics review board at the Hospital for Sick Children. The MDIG Trials was registered with clinicaltrials.org, with trial registration number: NCT01924013 (https://www.clinicaltrials.gov/ct2/show/NCT01924013).

### Blood Collection

Maternal and venous cord blood samples were collected at delivery by trained phlebotomists. Trace metal-free ethylenediaminetetraacetic acid (EDTA)-coated tubes were used to draw maternal samples and whole-blood aliquots were drawn prior to centrifugation. Cord blood was collected within 30 min of delivery from a site on the umbilical cord attached to the placenta after wiping maternal blood away with dry gauze. The umbilical vein was cannulated to collect blood into a lavender top EDTA tube. Whole blood was stored at -70ºC or colder.

### Measurement of selenium biomarkers

Whole blood and serum selenium were measured at the US Centers for Disease Control & Prevention (CDC). Whole blood selenium was measured at delivery and in venous cord blood using inductively coupled plasma mass spectrometry (ICP-MS).^22^ The lower limit of detection for this assay was 24.5 μg/L. Serum selenium was measured at delivery using ICP-dynamic reaction cell-MS (ICP-DRC-MS) as described by Caldwell (2011-2012).^23^ The lower limit of detection was 4.5 μg/L. No dilution factors were applied for these assays. Plasma SEPP1 was measured at delivery and in venous cord blood using a Selenotest ELISA colorimetric assay (STE, InVivo BioTech Services, Germany) performed at the Hospital for Sick Children. The lower limit of detection for the assay was 267.3 μg/L, with a 33-fold dilution factor applied and intra and inter-assay coefficients of variation (CVs) measured at 4.5 and 10.1% respectively. At least one selenium biomarker was measured in a sub-sample of 1021 specimens collected from mothers or infants for inclusion in this study.^21^

### Outcome Assessment

Details of outcome assessment have been elaborated on elsewhere.^20^ In brief, head-circumference, length, and weight were measured by trained professionals using standardized procedures adapted from the INTERGROWTH-21^st^ (International Fetal and Newborn Growth Consortium for the 21^st^ Century) Project.^24^ Infant length was measured using a ShorrBoard. Infant weight was measured using a digital scale. Length and weight were measured at birth, during visits at 1-2 months, 3, 6, 9, 12, 15, 21 and 24 months of age. For the purposes of our study, growth outcomes were assessed at birth, 12 and 24 months of age.

Two independent personnel measured each child and repeated measurements were taken. Measurements that differed between the two personnel by 7 mm for length or 50 g for weight were taken again. Means of the final pairs were used in further analysis. Interrater reliability was high. Length, weight and weight-for-length, body mass index (BMI), and head circumference were converted to age standardized z-scores using INTERGROWTH-21^st^ standards for newborn size, postnatal growth standards for preterm infants to 64-weeks of postmenstrual age (weight, length and head circumference only) or World Health Organization (WHO) child growth standards, as appropriate.^24^ All outcome variables were modelled continuously. We also examined three birth outcomes, namely LBW, classified as birth weight < 2500 grams, and SGA, defined as birthweight below the 10^th^ percentile of gestational age- and sex-matched healthy reference population, as dichotomous outcomes.^25^ Preterm birth (PTB) was defined as gestational age at birth less than 37 weeks.^26^

### Covariates

Covariates to be adjusted for in models included infant characteristics: sex, gestational age at birth and season of birth; maternal characteristics included age, education, height, body-mass index (BMI), gravidity, vitamin D treatment group, daily protein intake (kg), urinary cotinine levels, maternal smoking and tobacco use during pregnancy, C-reactive protein (CRP) concentrations at delivery, and household asset index.

Gestational age was estimated at time of enrollment using a combination of the last menstrual period and ultrasound. Smoking status was described using urinary cotinine levels, where women with cotinine ≤ 50 ng/mL were classified as non-smokers and women with levels > 50 ng/mL were classified as smokers.^27^ An asset index was derived using principal components analysis on baseline household characteristic data collected on ownership of various assets, where each participant was assigned an asset score (lower scores reflect less wealth).^20^ The asset index was categorized into quintiles.

### Statistical Analysis

All analyses were conducted using SAS 9.4 (SAS Institute, Cary, NC). A p-value < 0.05 was considered statistically significant. Descriptive statistics were generated for all outcome variables (growth indices) and selenium biomarkers as mean (± SD), median (IQR) depending on the shape of variable distributions. Bivariate correlations between all Se biomarkers were evaluated using Spearman correlation coefficients.

We assessed normality of all variables in this analysis using histograms and kernel density plots in order to determine their distribution. Outliers were flagged and examined to determine their biological plausibility, using box plots. Data for this secondary analysis were restricted to those participants with any one of the Se biomarkers in maternal delivery or venous cord specimens for which a growth measure was also available, as presented in **Figure 1**.

**Figure 1:**
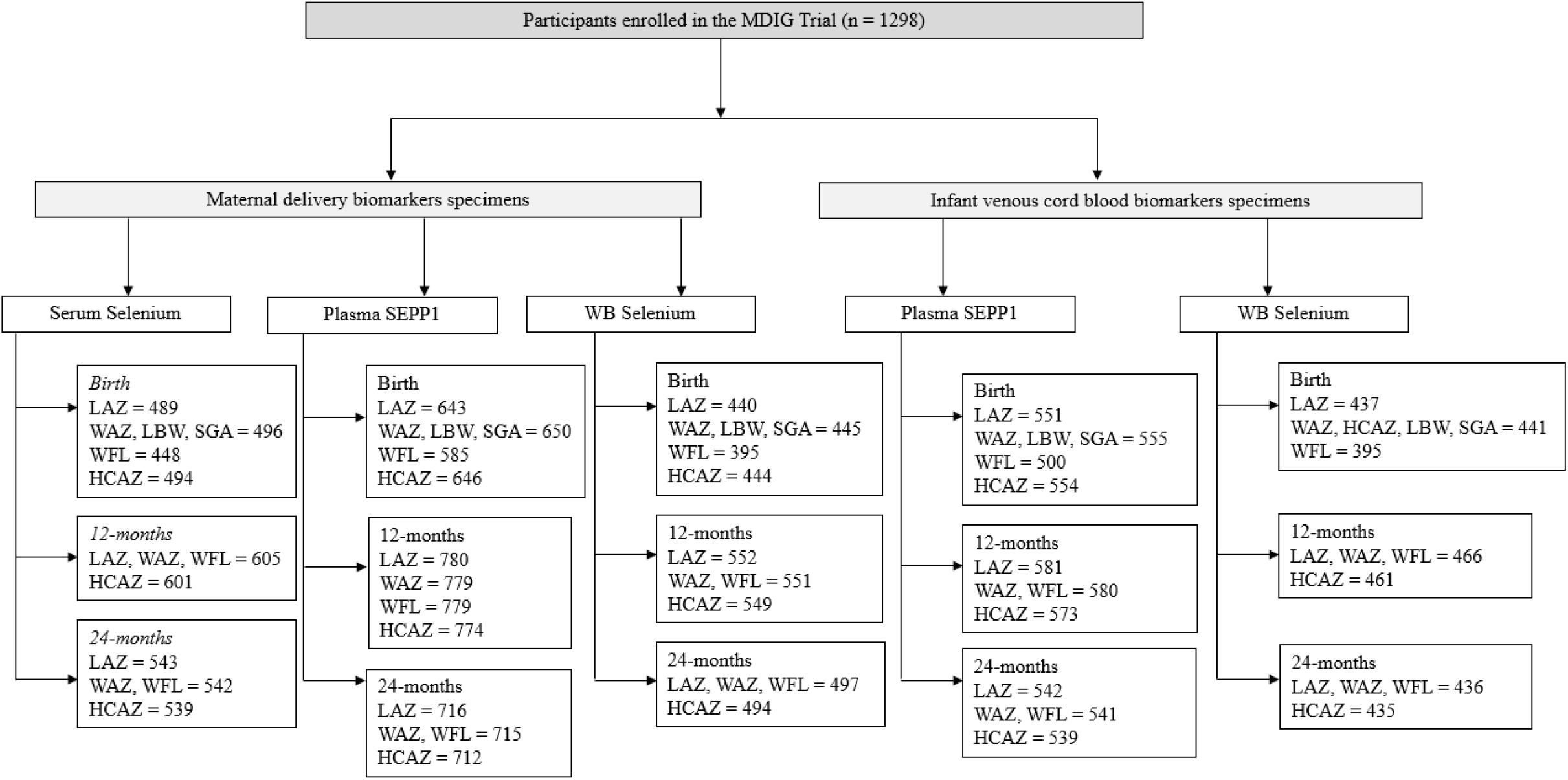
Sample size chart for maternal and cord selenium biomarkers included in analysis of associations with growth outcomes ^1^Length-for-age (LAZ); weight-for-age (WAZ); weight-for-length (WFL); head-circumference for age z-score (HCAZ) ^2^Low birth weight (LBW); small for gestational age (SGA)

We used linear regression and generated bivariate and multivariable models adjusting for covariates after generating directed acyclic graphs (DAGs) to account for hypothesized confounding and biological plausibility of associations. Biomarkers were scaled by a unit of 10 for whole blood and serum Se and unit of 1000 for SEPP1. Sensitivity analyses were conducted after including urinary cotinine as a covariate in adjusted models.

We also examined associations between selenoproteins GPx (glutathione peroxidase) and TrxR (thioredoxin reductase) measured in maternal plasma at mid-gestation (17-24 weeks) and delivery and growth outcomes using linear regression, associations with birth weight, specifically (presented in **Supplementary Tables 1 and 2**). Multicollinearity between predictors and covariates in models was assessed using variance inflation factors (VIFs), tolerance and collinearity using ‘proc reg’.

#### Se Biomarkers and Birth Outcomes

Descriptive statistics for n (%) of children who were SGA, LBW, and PTB in this sample were also examined. We conducted additional analyses to examine associations between SGA, LBW and PTB as dichotomous outcomes using modified Poisson regression with robust standard errors to estimate risk ratios (RRs) for associations between birth outcomes and selenium biomarker concentrations in unadjusted and adjusted models.

#### Mediation Analysis

In addition to linear models for growth outcomes, we conducted mediation analyses using observed variables to explore potential causal pathways for associations between cord whole blood Se and SEPP1 and weight at birth (birth weight and weight-for-age z-scores [WAZ]) and 1-year of age. We hypothesized that SEPP1 may mediate the association between WBSe in venous cord blood and WAZ/birth weight. We modelled both endogenous (outcome) and exogenous (predictor) variables continuously in structural equation modelling (SEM) path analysis.^28^ Mediation path analyses were conducted using ‘proc calis’ in SAS 9.4 and verified in RStudio using the ‘sem’ function. We examined direct, indirect, and total effects. Model fit was assessed using fit statistics including SRMR (< 0.08), RMSEA (< 0.08), χ^2^ (p-value > 0.05), CFI (≥ 0.90), GFI (≥ 0.95) and TLI (≥ 0.95) and benchmarked based on established criterion.^29^ We did not adjust for covariates in path analyses and results are presented as standardized estimates.

#### Longitudinal Models & Generalized Additive Models

We applied a gate-keeper approach to assess longitudinal associations between growth outcomes and selenium biomarkers based on significant associations observed in cross-sectional linear analyses at ages 0, 1 and 2-years. We used unadjusted multilevel models of change (‘proc mixed’) to assess whether relevant selenium biomarkers in venous cord blood predicted a change in growth-z-scores between 0-3 years based on availability of anthropometric data. We also used generalized additive models (‘proc gam’) to examine smoothing splines for predicted associations between selenium biomarkers and relevant measures of growth.

## Results

### Participant Characteristics

A detailed description of analytical sample sizes across growth outcomes in our study of mother-infant dyads is presented in **Figure 1**, for three timepoints, namely, birth, 12-months and 24-months of age. Given the distribution of observations with available Se biomarkers, our sample sizes ranged between 435-780 for these analyses across growth outcomes. On average, mothers were 23 years of age (SD ± 4), a majority were educated at a secondary school level or higher, median gestational age at birth was 39 weeks (IQR: 2) and mean ± SD birth weight across our sample was 2724 ± 354 grams, with close to 46% of children categorized as SGA, approximately 25% were LBW (< 2500 grams) and 8% were born before 37 weeks of gestation (**Table 1**). A small proportion of our sample was below the threshold (< 100 μg/L) for WBSe in maternal delivery and VC specimens (2.6% and 0.2%, respectively), and 83.4% of specimens were below threshold for serum Se at delivery (< 80 μg/L) and 0.2% of the sample was classified as deficient (< 30 μg/L). Biomonitoring equivalents for toxic concentrations of Se in a variety of body fluids have been calibrated using various regulatory limits set by the Institute of Medicine (IOM) (upper limits, UL), USEPA (reference dose, RfD) and ATSDR chronic (minimum risk level, MRL), and were established using Chinese cohort data to define guideline levels to protect against selenosis.^30^ These values range between 400-480 μg/L in whole blood, and 180-230 μg/L in plasma.^30^ In our sample, we found maximum levels of WBSe to range between 190.6-233.4 μg/L (∼50% less than WB ranges) in delivery and venous cord blood and 110.3 μg/L (∼ 60% less than WB ranges) in serum specimens, which do not suggest risk for toxicity due to selenosis in this population. Selenium biomarker concentrations are presented in **Table 2**. Spearman correlation coefficients for associations between delivery and venous cord selenium biomarkers in our analyses have been described in **Table 3**. We found small to moderate correlations between selenium biomarkers at delivery and in venous cord blood, with the exception of associations between SEPP1 and whole blood selenium in both specimen types.

**Table 1:** Descriptive characteristics of maternal and infant biomarkers of selenium (Se) status in the growth study

| **Maternal & Household Characteristics<sup>1,2</sup>** | **Maternal Delivery** |  |  | **Infant Venous Cord** |  |
| --- | --- | --- | --- | --- | --- |
|  | **Whole blood Se** | **Serum Se** | **SEPP1** | **Whole blood Se** | **SEPP1** |
| *N* | 467 | 517 | 677 | 448 | 562 |
| **Age (years), mean (SD)** | 23 (4) | 23 (4) | 23 (4) | 23 (4) | 23 (4) |
| **Gravidity, mean (SD)** | 2 (1) | 2 (1) | 2 (1) | 2 (1) | 2 (1) |
| **Height (cm), mean (SD)** | 151 (6) | 151 (5) | 151 (5) | 151 (6) | 151 (6) |
| **Gestational age at delivery (in weeks), median (IQR)** | 39.1 (1.7) | 39.1 (1.9) | 39.1 (1.7) | 39.1 (1.7) | 39.3 (1.9) |
| **Weight at delivery (kg), mean (SD)** | 61 (10) | 61 (10) | 61 (10) | 61 (10) | 61 (10) |
| **BMI (kg/m<sup>2</sup>), mean (SD)** | 24 (4) | 24 (4) | 24 (4) | 24 (4) | 24 (4) |
| **Ever smoked during pregnancy, n (%)** | 4 (0.9) | 3 (0.6) | 4 (0.6) | 2 (0.5) | 4 (0.7) |
| **Ever used tobacco during pregnancy, n (%)** | 179 (38) | 191 (37) | 235 (35) | 161 (36) | 195 (35) |
| **Urinary cotinine, n (%), (> 50 ng/mL)** | 18 (4) | 16 (3) | 16 (3) | 13 (3) | 13 (2) |
| **Daily protein intake (kg/day), mean (SD)** | 0.8 (0.3) | 0.9 (0.3) | 0.9 (0.4) | 0.9 (0.4) | 0.9 (0.4) |
| **Household asset index quintile, n (%)<sup>3</sup>** |  |  |  |  |  |
| 1 | 92 (20) | 94 (18) | 120 (18) | 85 (19) | 108 (19) |
| 2 | 94 (20) | 99 (19) | 132 (20) | 81 (18) | 102 (18) |
| 3 | 95 (20) | 115 (22) | 144 (21) | 103 (23) | 123 (22) |
| 4 | 96 (21) | 103 (20) | 139 (21) | 92 (21) | 113 (20) |
| 5 | 88 (19) | 102 (20) | 139 (21) | 84 (19) | 113 (20) |
| **Maternal Education, n (%)<sup>2</sup>** |  |  |  |  |  |
| *No education* | 20 (4) | 25 (5) | 27 (4) | 13 (3) | 19 (3) |
| *Primary incomplete* | 96 (21) | 104 (20) | 127 (19) | 88 (20) | 105 (19) |
| *Primary complete* | 64 (14) | 67 (13) | 93 (14) | 61 (14) | 76 (14) |
| *Secondary incomplete* | 175 (37) | 194 (38) | 267 (39) | 168 (38) | 214 (38) |
| *Secondary complete* | 112 (24) | 127 (25) | 163 (24) | 118 (26) | 148 (26) |
| **Season, n (%)<sup>2</sup>** |  |  |  |  |  |
| *Spring* | - | 67 (13) | 107 (16) | - | 88 (16) |
| *Summer* | 157 (35) | 160 (32) | 183 (28) | 168 (38) | 172 (31) |
| *Fall* | 175 (39) | 173 (34) | 220 (33) | 168 (38) | 182 (32) |
| *Winter* | 120 (27) | 106 (21) | 151 (23) | 111 (25) | 119 (21) |
| **Infant Characteristics<sup>3</sup>** |  |  |  |  |  |
| Sex (male), n (%) | 221 (49) | 236 (48) | 314 (48) | 224 (51) | 277 (50) |
| Gestational age at birth (weeks), *N* | 452 | 506 | 661 | 447 | 561 |
| median (IQR) | 39 (2) | 39 (2) | 39 (2) | 39 (2) | 39 (2) |
| **Birth Outcomes** |  |  |  |  |  |
| *N* | 445 | 496 | 650 | 441 | 555 |
| Birth weight (g) | 2706 (330) | 2721 (346) | 2721 (350) | 2742 (328) | 2747 (339) |
| Small-for-gestational age, n (%) | 224 (50) | 233 (47) | 302 (46) | 202 (46) | 253 (46) |
| Low birth weight (< 2500 g), n (%) | 124 (28) | 131 (26) | 169 (26) | 105 (24) | 130 (23) |
| Preterm birth (gestational age < 37 weeks), n (%)<sup>1</sup> | 39 (8) | 53 (10) | 64 (9) | 26 (6) | 33 (6) |
| *N* | 440 | 489 | 643 | 437 | 551 |
| Length-for-age z-score<sup>2, 4</sup> | -0.97 (0.99) | -0.88 (0.98) | -0.88 (0.99) | -0.88 (0.99) | -0.86 (1.00) |
| *N* | 445 | 496 | 650 | 441 | 555 |
| Weight-for-age z-score<sup>2, 4</sup> | -1.24 (0.82) | -1.17 (0.84) | -1.19 (0.86) | -1.14 (0.81) | -1.15 (0.83) |
| *N* | 395 | 448 | 585 | 395 | 500 |
| Weight-for-length z-score<sup>2, 4</sup> | -0.73 (0.94) | -0.72 (0.93) | -0.73 (0.95) | -0.65 (0.92) | -0.67 (0.93) |
| *N* | 444 | 494 | 646 | 441 | 554 |
| Head circumference-for-age z-score<sup>2, 4</sup> | -0.67 (0.94) | -0.62 (0.94) | -0.62 (0.95) | -0.59 (0.92) | -0.59 (0.94) |
| **12-months Growth Outcomes** |  |  |  |  |  |
| *N* | 552 | 605 | 780 | 466 | 581 |
| Length-for-age z-score<sup>2,4</sup> | -1.01 (1.01) | -0.99 (1.04) | -0.99 (1.06) | -0.89 (1.00) | -0.87 (1.02) |
| *N* | 551 | 605 | 779 | 466 | 580 |
| Weight-for-age z-score<sup>2,4</sup> | -0.88 (1.06) | -0.88 (1.09) | -0.87 (1.10) | -0.74 (1.08) | -0.78 (1.10) |
| *N* | 551 | 605 | 779 | 466 | 580 |
| Weight-for-length z-score<sup>2,4</sup> | -0.52 (1.01) | -0.53 (1.03) | -0.52 (1.04) | -0.42 (1.04) | -0.48 (1.06) |
| *N* | 549 | 601 | 774 | 461 | 573 |
| Head circumference-for-age z-score<sup>2,4</sup> | -1.23 (0.96) | -1.18 (0.99) | -1.19 (0.98) | -1.14 (0.90) | -1.16 (0.99) |
| **24-months Growth Outcomes** |  |  |  |  |  |
| *N* | 497 | 543 | 716 | 436 | 542 |
| Length-for-age z-score<sup>2,4</sup> | -1.28 (1.04) | -1.32 (1.08) | -1.32 (1.09) | -1.22 (1.05) | -1.22 (1.07) |
| *N* | 497 | 542 | 715 | 436 | 541 |
| Weight-for-age z-score<sup>2,4</sup> | -1.17 (1.16) | -1.21 (1.17) | -1.20 (1.16) | -1.07 (1.18) | -1.11 (1.20) |
| *N* | 497 | 542 | 715 | 436 | 541 |
| Weight-for-length z-score<sup>2,4</sup> | -0.72 (1.09) | -0.74 (1.09) | -0.71 (1.07) | -0.61 (1.12) | -0.67 (1.14) |
| *N* | 497 | 539 | 712 | 435 | 539 |
| Head circumference-for-age z-score<sup>2,4</sup> | -1.30 (0.94) | -1.28 (0.97) | -1.30 (0.96) | -1.25 (0.88) | -1.28 (0.99) |

| **Vitamin-D Treatment Group** |  |  |  |  |  |
| --- | --- | --- | --- | --- | --- |
| Placebo control, n (%) | 133 (17) | 67 (15) | 2171 (788) | 155 (21) | 1481 (486) |
| 4,200 IU/week, n (%) | 133 (17) | 69 (11) | 2162 (761) | 155 (24) | 1472 (496) |
| 16,800 IU/week, n (%) | 136 (20) | 70 (12) | 2161 (666) | 161 (20) | 1597 (592) |
| 28,000 IU/week pregnancy only, n (%) | 135 (19) | 67 (14) | 2087 (740) | 163 (22) | 1550 (461) |
| 28,000 IU/week pregnancy & postpartum, n (%) | 131 (15) | 69 (12) | 2138 (605) | 162 (22) | 1566 (553) |

<sup>1</sup>Whole blood Se < threshold (< 100 µg/L), serum Se < threshold (< 80 µg/L), serum Se deficiency (< 30 µg/L), no thresholds exist for SEPP1
<sup>2</sup>Biomarker concentrations are un-scaled, sample sizes represent age at birth (up to 1-day postpartum) and maximum observations with Se biomarkers available
<sup>3</sup>Sample sizes do not correspond to totals for biomarker concentrations, observations without corresponding Se biomarkers are excluded
<sup>4</sup>All values are presented as mean (SD) unless indicated otherwise

**Table 2:**
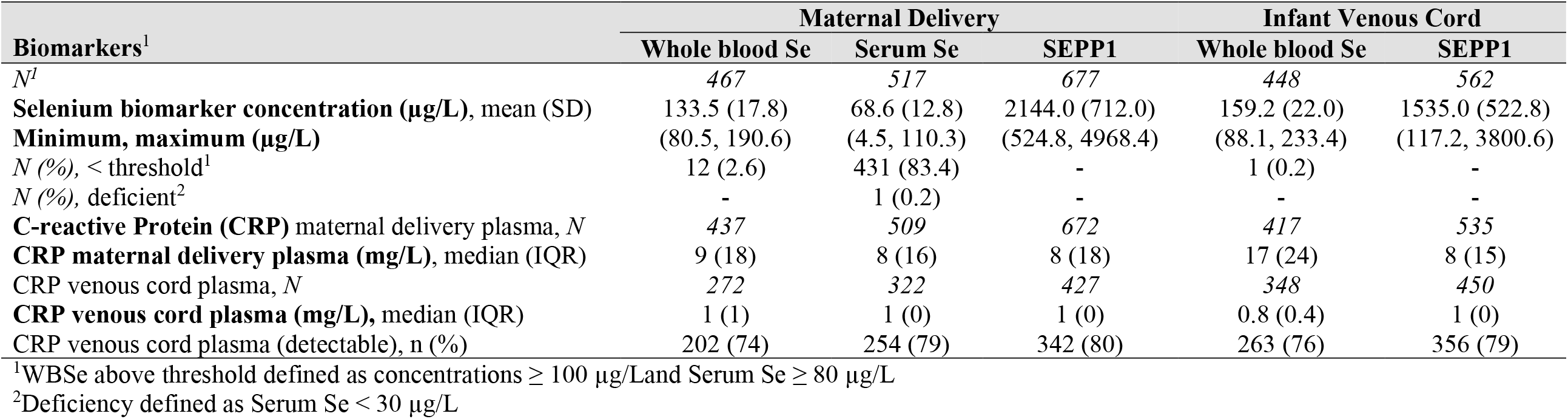
Selenium biomarker concentrations at delivery and in infant venous cord blood

| **Biomarkers<sup>1</sup>** | **Maternal Delivery** |  |  | **Infant Venous Cord** |  |
| --- | --- | --- | --- | --- | --- |
|  | **Whole blood Se** | **Serum Se** | **SEPP1** | **Whole blood Se** | **SEPP1** |
| *N*<sup>1</sup> | 467 | 517 | 677 | 448 | 562 |
| **Selenium biomarker concentration (µg/L), mean (SD)** | 133.5 (17.8) | 68.6 (12.8) | 2144.0 (712.0) | 159.2 (22.0) | 1535.0 (522.8) |
| **Minimum, maximum (µg/L)** | (80.5, 190.6) | (4.5, 110.3) | (524.8, 4968.4) | (88.1, 233.4) | (117.2, 3800.6) |
| *N* (%), < threshold<sup>1</sup> | 12 (2.6) | 431 (83.4) | - | 1 (0.2) | - |
| *N* (%), deficient<sup>2</sup> | - | 1 (0.2) | - | - | - |
| **C-reactive Protein (CRP) maternal delivery plasma, *N*** | 437 | 509 | 672 | 417 | 535 |
| **CRP maternal delivery plasma (mg/L), median (IQR)** | 9 (18) | 8 (16) | 8 (18) | 17 (24) | 8 (15) |
| **CRP venous cord plasma, *N*** | 272 | 322 | 427 | 348 | 450 |
| **CRP venous cord plasma (mg/L), median (IQR)** | 1 (1) | 1 (0) | 1 (0) | 0.8 (0.4) | 1 (0) |
| **CRP venous cord plasma (detectable), *n* (%)** | 202 (74) | 254 (79) | 342 (80) | 263 (76) | 356 (79) |

<sup>1</sup>WbSe above threshold defined as concentrations $\geq 100$ µg/L and Serum Se $\geq 80$ µg/L
<sup>2</sup>Deficiency defined as Serum Se < 30 µg/L

**Table 3:** Spearman correlation coefficients (*ρ*) for maternal delivery and venous cord selenium biomarkers

|  | Maternal Delivery |  |  |  |  |  |  |  |  | Infant Venous Cord |  |  |  |  |  |
| --- | --- | --- | --- | --- | --- | --- | --- | --- | --- | --- | --- | --- | --- | --- | --- |
|  | WBS<sub>e</sub> |  |  | Serum Se |  |  | SEPP1 |  |  | WBS<sub>e</sub> |  |  | SEPP1 |  |  |
| | n | $\rho$ | p-value | n | $\rho$ | p-value | n | $\rho$ | p-value | n | $\rho$ | p-value | n | $\rho$ | p-value |
| WBS<sub>e</sub> Del | 447 | 1 | - | 376 | 0.6 | <0.0001 | 385 | 0.3 | <0.0001 | 332 | 0.5 | <0.0001 | 320 | -0.00001 | 0.9 |
| Serum Se Del | 376 | 0.6 | <0.0001 | 497 | 1 | - | 487 | 0.4 | <0.0001 | 321 | 0.3 | <0.0001 | 389 | 0.1 | 0.02 |
| SEPP1 Del | 385 | 0.3 | <0.0001 | 487 | 0.4 | <0.0001 | 651 | 1 | - | 377 | 0.03 | 0.6 | 488 | 0.2 | <0.0001 |
| WBS<sub>e</sub> VC | 332 | 0.5 | <0.0001 | 321 | 0.3 | <0.0001 | 377 | 0.03 | 0.6 | 443 | 1 | - | 404 | 0.1 | 0.001 |
| SEPP1 VC | 320 | -0.00001 | 0.9 | 389 | 0.1 | 0.02 | 488 | 0.2 | <0.0001 | 404 | 0.2 | 0.001 | 557 | 1 | - |

### Modelling Analyses

Linear regression analyses are presented for unadjusted and adjusted associations between selenium biomarkers and growth outcomes in **Table 4**.

**Table 4:**
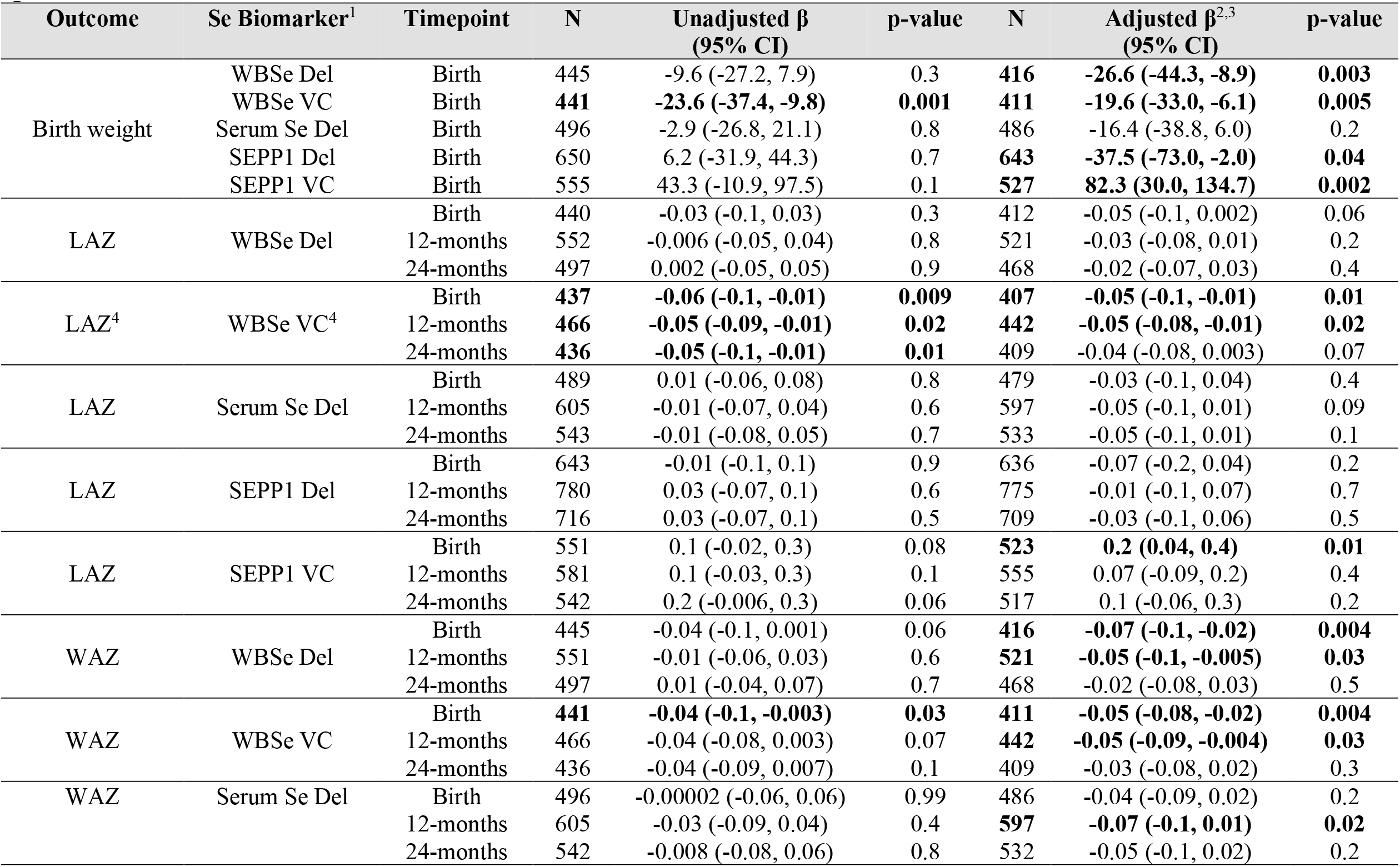
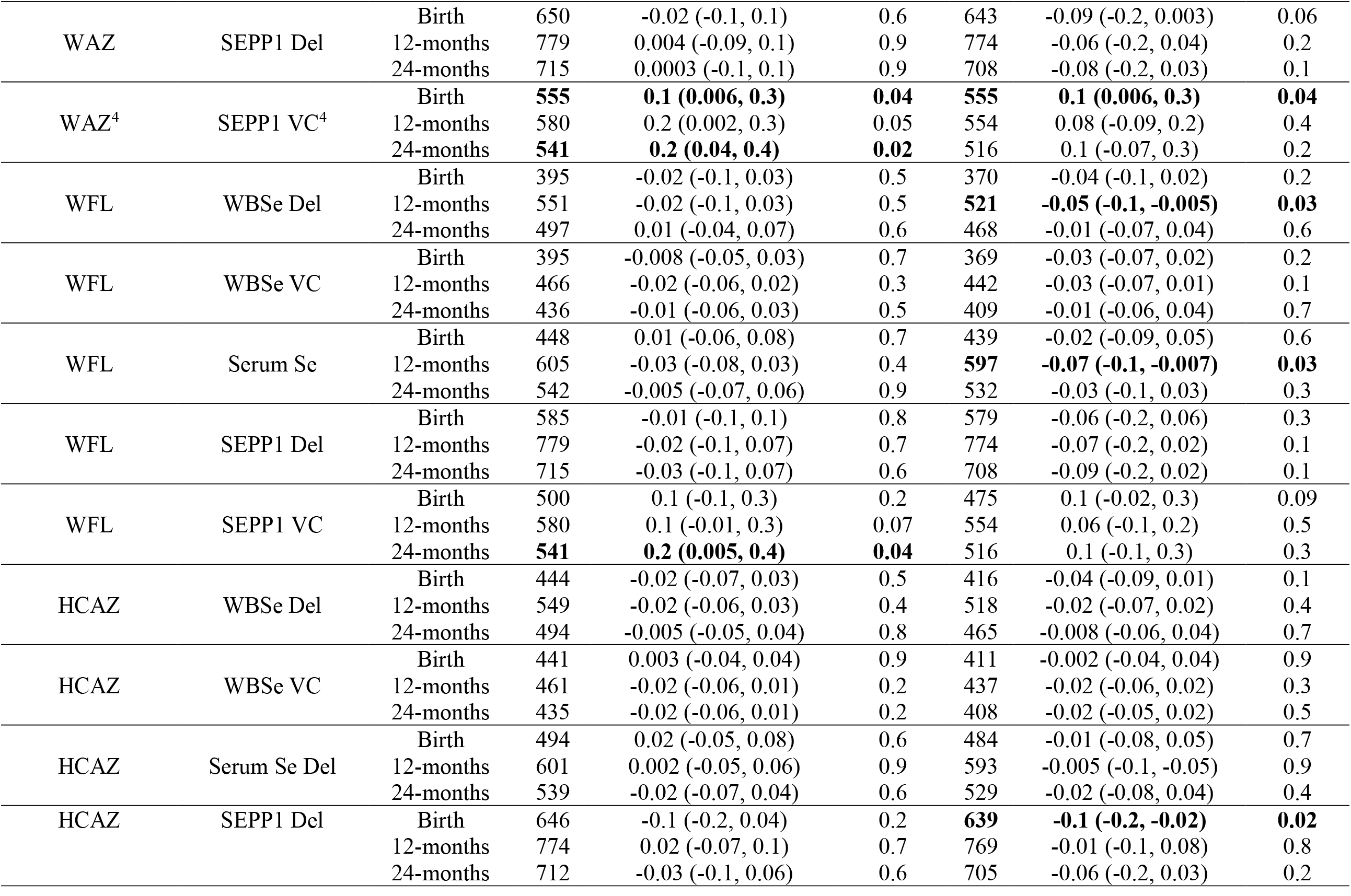
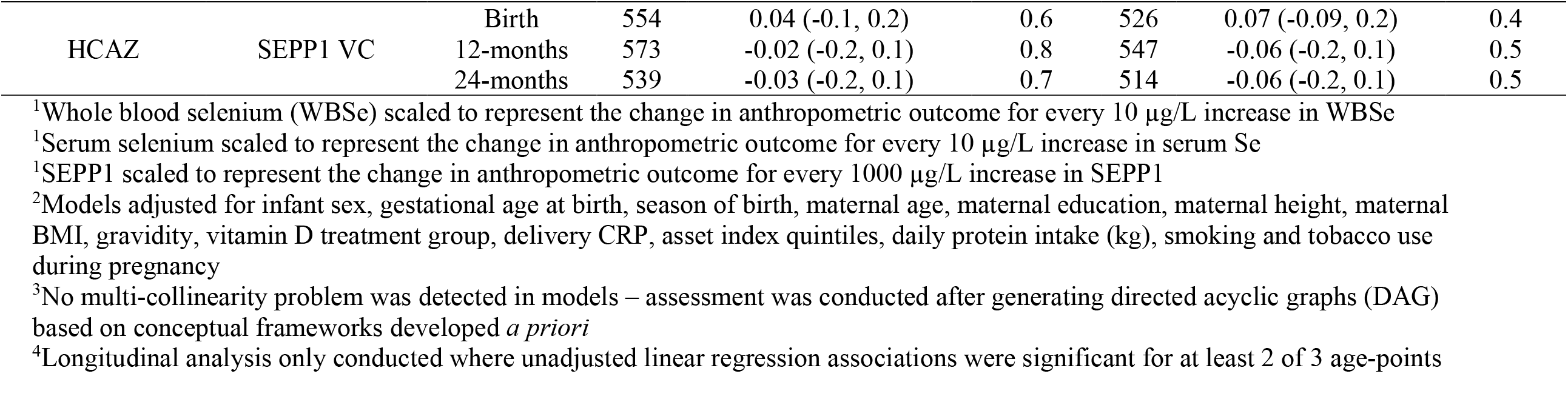
Linear and longitudinal regression analyses for associations between maternal delivery and venous cord Selenium biomarkers and growth outcomes

#### Whole blood selenium and growth outcomes

We examined associations between whole blood selenium and birth weight among infants in our sample. We found negative associations between delivery WBSe and birth weight in unadjusted models, where per scaled unit increase in WBSe, birth weight decreased by 10-grams.

Likewise, we found null associations (RR: 1.03, 95% CI: 0.96 to 1.1) for LBW per unit (gram) increase in maternal delivery WBSe in unadjusted models, with increased risk (12-15%) after accounting for confounders.

In line with our findings for maternal delivery WBSe, we found estimates for associations between venous cord WBSe and birth weight to be significant with effect estimates suggesting a decrease in birthweight of between 24-27 grams per unit increase in selenium, between unadjusted and adjusted models. Results for risk of LBW were in concordance with our findings for birth weight, with a 6-11% increased risk of the outcome per unit increase in WBSe in cord blood as presented in **Table 5**. We found a non-significant yet increased risk of SGA status per unit increase in WBSe in maternal blood at delivery and in venous cord blood. Similarly, we found higher risk for SGA with adjusted models showing significant RRs (7-10% increase per unit increase in venous cord WBSe).

**Table 5:** Modified Poisson Regression analyses for associations between maternal delivery and venous cord selenium biomarker and birth outcomes

| Outcome | Se Biomarker<sup>1</sup> | Timepoint | N | Unadjusted Risk Ratio<br>(95% CI) | p-value | N | Adjusted Risk Ratio<sup>2</sup><br>(95% CI) | p-value |
| --- | --- | --- | --- | --- | --- | --- | --- | --- |
| Small-for-gestational age (SGA) | WBSe Del | Delivery | 445 | 1.03 (0.96, 1.1) | 0.4 | 416 | 1.04 (0.96, 1.1) | 0.3 |
|  | WBSe VC | Venous Cord | 441 | 1.06 (0.99, 1.1) | 0.07 | 411 | 1.07 (1.0, 1.1) | 0.05 |
|  | Serum Se Del | Delivery | 496 | 1.02 (0.9, 1.1) | 0.7 | 486 | 1.05 (0.9, 1.2) | 0.4 |
|  | SEPP1 Del | Delivery | 650 | 0.99 (0.85, 1.2) | 0.9 | 643 | 1.03 (0.9, 1.2) | 0.7 |
|  | SEPP1 VC | Venous Cord | 555 | 0.8 (0.6, 1.0) | 0.08 | 527 | 0.7 (0.5, 0.9) | 0.01 |
| Low birth weight (LBW) | WBSe Del | Delivery | 445 | 1.05 (0.95, 1.2) | 0.3 | 416 | 1.12 (1.0, 1.3) | 0.04 |
|  | WBSe VC | Venous Cord | 441 | 1.11 (1.0, 1.2) | 0.02 | 411 | 1.08 (0.99, 1.2) | 0.1 |
|  | Serum Se Del | Delivery | 496 | 1.06 (0.93, 1.2) | 0.4 | 486 | 1.12 (0.97, 1.3) | 0.1 |
|  | SEPP1 Del | Delivery | 650 | 0.99 (0.85, 1.2) | 0.9 | 643 | 1.2 (1.0, 1.5) | 0.1 |
|  | SEPP1 VC | Venous Cord | 555 | 0.8 (0.6, 1.0) | 0.08 | 527 | 0.9 (0.6, 1.3) | 0.5 |
| Preterm birth (PTB) | WBSe Del | Delivery | 467 | 1.01 (0.8, 1.2) | 0.9 | 418 | 1.31 (0.9, 1.9) | 0.2 |
|  | WBSe VC | Venous Cord | 448 | 1.13 (0.96, 1.3) | 0.2 | 413 | 1.01 (0.8, 1.3) | 0.9 |
|  | Serum Se Del | Delivery | 517 | 1.11 (0.9, 1.4) | 0.3 | 487 | 0.99 (0.7, 1.4) | 0.9 |
|  | SEPP1 Del | Delivery | 677 | 0.95 (0.7, 1.3) | 0.8 | 644 | 1.06 (0.67, 1.7) | 0.8 |
|  | SEPP1 VC | Venous Cord | 562 | 0.5 (0.26, 1.0) | 0.06 | 529 | 1.2 (0.5, 3.0) | 0.7 |

<sup>1</sup>Whole blood selenium (WBSe) scaled to represent the change in anthropometric outcome for every 10 µg/L increase in WBSe
<sup>1</sup>Serum selenium scaled to represent the change in anthropometric outcome for every 10 µg/L increase in serum Se
<sup>1</sup>SEPP1 scaled to represent the change in anthropometric outcome for every 1000 µg/L increase in SEPP1
<sup>2</sup>Models adjusted for infant sex, season of birth, maternal age, maternal education, maternal height, maternal BMI, gravidity, vitamin D treatment group, delivery CRP, asset index quintiles, daily protein intake (kg), smoking and tobacco use during pregnancy

Associations between LAZ and WBSe in maternal delivery samples were not significant, although we found small negative associations between WBSe in venous cord blood and LAZ at birth, 12 and 24-months of age. Although effect estimates for associations between WBSe in maternal delivery samples and LAZ were small and in the negative direction, we did not find statistical significance across analyses for WBSe and growth outcomes across age time points. For associations between WBSe in venous cord blood and LAZ, we found small negative effect estimates across unadjusted models at birth, 1-year, and 2-years of age. Although mildly attenuated after adjustment, associations between WBSe in venous cord blood and LAZ were no longer significant, or in sensitivity analyses, after adjusting for cotinine **(Supplementary Tables 3 and 4)**. In longitudinal analyses, we found mean LAZ at birth to be -0.12 (SE: 0.27), p = 0.27, not significantly different from 0. Average change in LAZ as children age, per unit increase in venous cord WBSe was -0.004 (SE: 0.0017), p = 0.019 (AIC: 7392.3, BIC: 7416.2), which suggests that this form of selenium has a marginal and non-significant association with linear growth.

Associations between WAZ and delivery whole blood selenium, although similar in magnitude to those observed for LAZ, were not significant in unadjusted models; however, effect estimates increased marginally, and significance was achieved after adjustment and in sensitivity analyses with cotinine. We found small negative associations between WAZ and WBSe in venous cord at birth in unadjusted and adjusted models. Overall trends ran in the same direction across unadjusted and adjusted models for other growth outcomes including weight-for-length and head circumference for age z-scores.

#### Serum Se and growth outcomes

In line with our observations for WBSe, we found similar trends for associations between serum selenium at delivery and growth measures, although associations were small and non-significant. Risk ratios were also higher for SGA and LBW per unit increase in serum selenium concentrations. Similar to results for WBSe, associations between serum selenium and birth weight were also negative, and large in magnitude (3-28 gram decrease in birthweight per unit increase in serum selenium), although these findings were not significant and confidence intervals for effect estimates were wide.

#### SEPP1 and growth outcomes

SEPP1, unlike serum and WBSe, showed positive associations with infant birth weight in unadjusted and adjusted models, although results for statistical significance were not consistent across models. Based on our findings, a per unit increase in venous cord blood SEPP1 was associated with a large increase in birthweight, with estimates ranging between 43-82 grams between unadjusted and adjusted models. Risk for LBW in our analyses was marginal, with WBSe in venous cord blood suggesting a protective effect per unit increase in concentrations, although results were non-significant.

We did not see significant or consistent associations between LAZ at any age time point and SEPP1 in maternal delivery plasma samples in unadjusted models. We did observe small positive increases in LAZ scores in adjusted models at birth when examining SEPP1 concentrations in venous cord blood, and associations were significant after accounting for covariates in adjusted models. Effect estimates suggest a stronger association between venous cord SEPP1 and LAZ at birth up to 2 years of age. These associations, although similar in magnitude were not significant at 12 months and 24 months of age.

Non-significant positive associations were observed between WAZ and SEPP1 in maternal delivery samples at birth. Significant associations were observed between SEPP1 in venous cord blood and WAZ. We used uncontrolled longitudinal hierarchical models and found a mean WAZ at birth to be -1.1 (SE: 0.1), p <0.001. Average change in WAZ as children age, per unit increase in VC SEPP1 was 0.0001 (SE: 0.0001), p <0.05 (AIC: 11047.9, BIC: 11083.5). These findings suggest a marginal protective effect of this form of selenium on WAZ.

WFL and HCAZ were not associated with SEPP1 in maternal delivery and/or venous cord blood. We did note that the overall risk for SGA and LBW was marginally lower per unit increase in SEPP1 in both maternal delivery and venous cord blood; however, these relationships were not significant. Across the different biomarkers, we did note shifts in the direction of trends across the different age points at which growth outcomes were measured.

#### Mediation analyses and Generalized Additive Model analyses

Results from mediation path analysis are presented in **Figures 2, 3** and **4**. Across path models, the indirect mediation effects, which represent the association of WBSe and weight-related growth outcomes, mediated through SEPP1, were extremely small and non-significant. We found small but significant direct effects between WBSe and SEPP1 in venous cord blood and growth outcome measures. Overall fit indices for all path models suggest statistical convergence and good fit.

**Figure 2:**
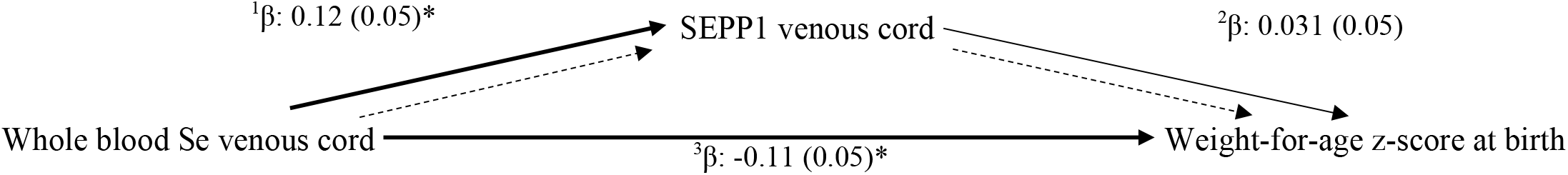
Mediation path analysis exploring associations between WAZ at birth and venous cord selenium biomarkers ^a,b^Unadjusted direct and indirect effects in mediation analysis (n = 402) ; Total effect: β: -0.11 (0.05)*; Indirect effect: β: 0.004 (0.006) ^c^Fit-indices for models: χ^2^: 11.3, df: 3, p-value: 0.01; CFI: 1.00; TLI: 1.00; SRMR: 0.00; GFI: 1.00; RMSEA: 0.000 (overall model fit is good) *p < 0.05, ** p < 0.01

**Figure 3:**
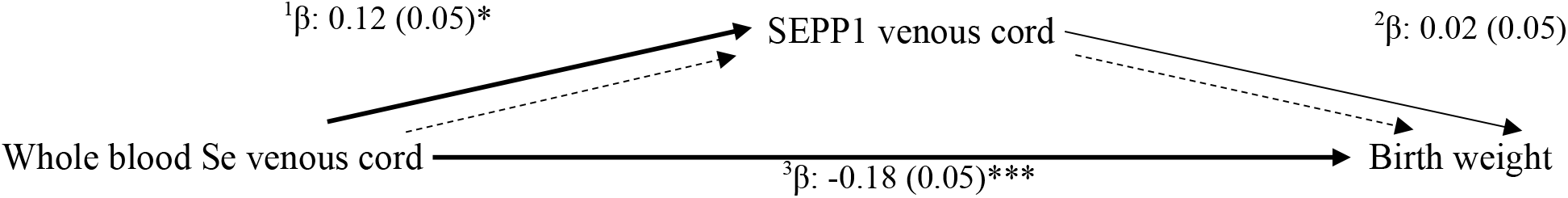
Mediation path analysis exploring associations between birthweight and venous cord selenium biomarkers ^a,b^Unadjusted direct and indirect effects in mediation analysis (n = 402) ; Total effect: β: -0.18 (0.05)***; Indirect effect: β: 0.003 (0.01) ^c^Fit-indices for models: χ^2^: 19.5, df: 3, p-value: 0.0002; CFI: 1.00; TLI: 1.00; SRMR: 0.00; GFI: 1.00; RMSEA: 0.000 (overall model fit is good) ^d^Unstandardized estimates for path models (to compare beta-estimates to linear regression models): ^1^β: 0.03 (0.01)*; ^2^β: 13.1 (29.8); ^3^β: -26.9 (7.3)** *p < 0.05, ** p < 0.01, ***p < 0.001, ****p < 0.0001

**Figure 4:**
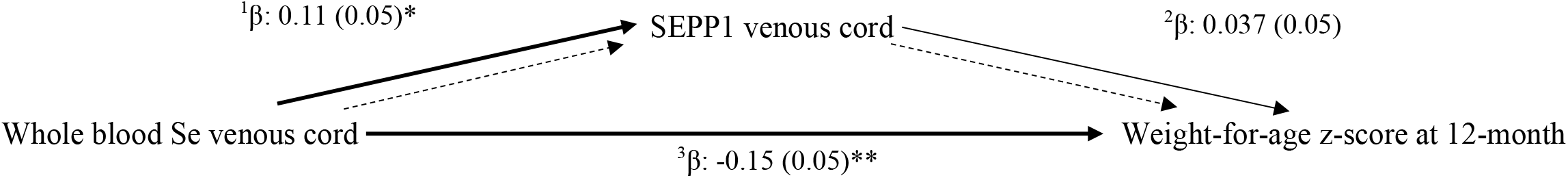
Mediation path analysis exploring associations between WAZ at 12-months of age and venous cord selenium biomarkers ^a,b^Unadjusted direct and indirect effects in mediation analysis (n = 394); Total effect: : β: -0.15 (0.05)**; Indirect effect: β: 0.004 (0.006) ^c^Fit-indices for models: χ^2^: 13.7, df: 3, p-value: 0.003; CFI: 0.00; TLI: 1.00; SRMR: 0.00; GFI: 1.00; RMSEA: 0.000 (overall model fit is good) *p < 0.05, **p < 0.01

To examine dose-response relationships between length-for-age and weight-for-age, we performed GAM analysis with Se biomarkers in maternal delivery specimens and venous cord blood. Figures are presented in **Supplementary Figures 1 and 2**. We found consistent inverse U-shaped associations between LAZ, WAZ and SEPP1 in venous cord blood and similarly between growth z-scores and serum selenium in delivery specimens.

## Discussion

In this study of mother-infant dyads, we found measures of selenium status in maternal delivery and venous cord blood serum, whole blood, and plasma SEPP1 to be modestly associated with infant growth outcomes at birth, 12 months, and 24 months of age, with variability in the strength and magnitude of associations across biomarkers and age points. Effect sizes in path analyses were negligible and do not suggest any mediation of the association between WBSe and WAZ at birth or 12-months, by venous cord SEPP1, likely because WBSe and SEPP1 have differing functions and therefore different impacts on weight and act as part of different metabolic pools that do not interact or share common pathways, as described in **Supplementary Figure 3**.^31^

Several studies have examined the effects of selenium in a variety of biological specimen types and their associations with infant growth outcomes.^12, 19, 32-42^ A comprehensive comparative list of epidemiological studies has been presented in **Table 6**. Overall concentrations of selenium across biomarker types in our study were comparable to those described in the literature. During pregnancy, Se progressively accumulates in fetal organs.^43^ Selenium is stored primarily in the liver of the growing fetus and is likely redistributed after birth, where concentrations in utero remain constant during gestation and decrease in line with stable selenium concentration in other organs, during the months following birth.^8^ Transfer of SEPP1 from maternal blood to fetus is facilitated by Apo-ER2 mediated endocytosis. Selenomethioneine containing proteins and SeMet also supply selenium to the fetus.^44-45^

**Table 6:** Epidemiological studies examining associations between Se status during pregnancy, postpartum and in cord blood and birth/growth outcomes

| Authors | Study Design & Sample Size | Measures of Se status | Measures of birth/growth outcomes | Key Findings |
| --- | --- | --- | --- | --- |
| Bogden et al., 2006 | CA: (<37 weeks gestation, n = 107; sub-set <32 wks, n = 25) | Maternal erythrocyte GPx activity (microunits of GPx activity/mg of HgB): 20.9±0.76; 22.4±0.7 | Serum Se & birth weight (g): -134±182 (p = 0.46) (lowest decile vs. others, CA, n = 126) | Low-normal serum Se measured in 1<sup>st</sup> and 2<sup>nd</sup> trimesters of pregnancy predict lower birth weight for full-term infants. The risk of LBW was highest in the lowest decile of serum Se, no increase in risk above lowest decile<br>Reduction in birth weight was considerable (-259 gms for women in lowest decile of serum Se) |
|  | CO: n = 126, ≥37 gestation) nested within cohort | Dietary Se intake (24-hr recall): 115±9; 93±5<br>Serum Se (umol/L): 1.37±0.02; 1.34±0.02<br>Collected in first and second trimesters of pregnancy | Serum Se & birth weight (g): -270±117 (p = 0.02) (lowest decile vs. others, CO, n = 107) |  |
| Klapec et al., 2008 | Cross-sectional mother newborn pairs<br>IUGR – n = 57;<br>normal – n = 44 | Placental Se levels (ug/g) | Placental level Se (ug/g) & IUGR (median, range): 0.14 (0.10, 0.20), n = 49; AGA (n = 36): 0.15 (0.10, 0.24) | Gestational age, Se content in placenta, maternal BMI, newborn sex were associated with significant contribution to birth weight in AGA infants |
|  |  |  | Se (ug/g) vs. weight change (g/unit): 6954.92 (508.25, 11125.61), p <0.05, R<sup>2</sup>: 0.59 | Maternal serum or placental Se – predict birth weight for full-term infants |
| Tsuzuki et al., 2013 | Japan, observational<br>44 infants<br>14 premature<br>2 IUGR | Maternal serum Se at delivery: (pre-term: 79.3±19.3 ug/L vs. term: 94.1±18.1 ug/L, p = 0.032)<br>Cord blood Se at delivery: (pre-mature: 52.8±9.7 ug/L vs. term: 59.4±10.0 ug/L, p = 0.071) | Birth weight (dependent):<br>NeoSe: 17.783 (-5.590, 41.155); p = 0.128<br>MatSe: 10.985 (2.391, 19.578); p = 0.015 (maternal [serum Se]) | Selenium status in neonates and mothers is positively associated with birth weight |
|  |  | Neonatal serum Se at day: (pre-term: 43.3±7.0 ug/L vs. term: 52.0±8.9 ug/L, p = 0.001) | predictor of neonatal birth weight independent of maternal nutritional status) |  |
|  |  |  | UmbSe: -14.524 (-33.134, 4.085); p = 0.119 |  |
| Mistry et al., 2014 | 126 pregnant adolescent women<br>Prospective observational study – About Teenage Eating Study | [Selenium ug/L]<br>AGA (n = 107) → 65.1 (62.7, 67.5)<br>SGA (n = 19) → 49.4 (45.9, 52.9) | SGA: individualized birth weight ratio < 10<sup>th</sup> percentile; plasma Se: 60.3 ug/L (White-European); 65.9 ug/L (Afro-Caribbean)<br>OR for SGA: 1.15 (1.07, 1.23); 1.16 (1.08, 1.24), p <0.0001 | Odds of SGA are significantly higher among pregnant women with lower plasma Se |
| Sun et al., 2014 | 209 women, cross-sectional study (19-41 years) | Arithmetic mean (95% CI)<br>Maternal Se: 156.58 (145.82, 167.34) ug/L<br>Cord Se: 8.71 (7.29, 10.14) ug/g<br>Urine Se: 129.04 (124.30, 133.76) ug/L | Log cord Se & birth weight: 653.80. p = 0.01 | Cord blood selenium is positively associated with birth weight |
| Nazemi et al., 2015 | Case: 91 mothers (LBW < 2500 gms)<br>Control: 86 mothers (normal birth weight neonates) | Maternal blood Se (ug/mL):<br>CA: 80.69±28; CO: 78.48±25.54<br>Umbilical blood Se (ug/mL):<br>CA: 77.32±26.12; CO: 73.89±24.37 | Maternal blood Se, p-value: 0.961<br>Umbilical cord Se, p-value: 0.660 | No significant associations found between maternal or umbilical cord Se and birth weight |
| Bermudez et al., 2015 | Cross-sectional, n = 54 paired samples<br>SGA, n = 11; AGA, n = 30; LGA, n = 13 | Cord plasma Se (nM/L):<br>438.4±78.6<br>Maternal plasma Se (nM/L):<br>704±199.7<br>Cord/maternal ratio (mean ± SD):<br>1.08±3.02 | Umbilical cord blood Se & SGA (n = 11): 442.0±79.81 nM/L<br>Umbilical cord blood Se & AGA (n = 30): 439.0±84.83 nM/L<br>Umbilical cord blood Se & LGA (n = 13): 433.8±67.37 nM/L | Umbilical cord selenium was higher in SGA neonates, however maternal plasma Se was lower in SGA infants, suggesting preferential transfer to the fetus to sustain growth and development |
|  |  |  | Maternal peripheral blood Se & SGA (n = 8): 680.9±298.6nM/L<br>Maternal peripheral blood Se & AGA (n = 25): 712.37±160.2 nM/L<br>Maternal peripheral blood Se & LGA (n = 13): 701.92±213.91 nM/L |  |
| Hu et al., 2016 | N = 81 mother-newborn pairs from 4 cities in China enrolled cross-sectionally | Maternal blood Se (ng/g): 140.8<br>Newborn cord blood Se (ng/g): 22.0 | Birth weight & maternal blood: $\beta$:1.0 (-3.6 to 1.6), p = 0.459<br>$\beta$:0.5 (-3.2 to 2.2), p = 0.707 | Positive non-significant associations observed between maternal selenium levels and infant birth weight |
| Kobayashi et al., 2019 | Japan Environmental & Children's Study – prospective birth cohort, n = 15,444 pregnant women | Blood Se: 170.0 (158.0-183.0) ng/g | Compared to infants of mothers with highest blood Se, mothers with lowest blood Se had neither significant increase in birth weight (9 g, 95% CI: -6.25) nor significant odds ratio of SGA (0.903, 95% CI: 0.748, 1.089) | No observed differences between birth weight and SGA and levels of maternal blood Se |
| Lewandowska et al., 2019 | Nested prospective cohort of n = 750 women recruited at 10-14<sup>th</sup> week of singleton healthy pregnancy in Poland | Mean Serum Se concentration of cohort: 61.95 ug/L (range: 41.14-89.17 ug/L) | SGA: (weight < 10<sup>th</sup> percentile for GA, gender of newborn in population): n = 48<br>AGA: (weight between 10-90<sup>th</sup> percentile): n = 192<br>Q<sub>1</sub>: Se: [41.14-56.60 ug/L]; OR for SGA: 2.63 (1.08-6.42); p = 0.034<br>Q<sub>2</sub>: Se: [56.60-61.86 ug/L]; OR for SGA: 0.87 (0.36-2.14); p = 0.794 | Lowest concentrations of early pregnancy maternal serum Se associated with higher risk of SGA<br>Threshold found at 56.1 ug/L below which steep increase in SGA risk observed |
|  |  |  | Q<sub>3</sub>: Se: [61.86-66.62 ug/L]; OR for SGA: 1.42 (0.55-3.66); p = 0.472<br>Q<sub>4</sub>: Se: [66.62-89.17 ug/L]; OR for SGA: 1 |  |
| Sole-Navais et al., 2020 | MoBa – prospective population-based pregnancy cohort, Norway | WbSe: (n = 2572 women at 17-18 weeks gestation), median: 102 (89-117) ug/L | Maternal dietary Se intake in first 4-5 months of pregnancy: $\beta$: 0.027 (95% CI: 0.007, 0.014)<br>Birth weight (gms): $\beta$: -0.47 (-1.29, 0.34), 41.6; p = 0.254<br>Z-scores: $\beta$: -0.001 (-0.003, 0.001), 0.09, p = 0.195<br>SGA OR: 1.24 (0.53, 2.88), p = 0.623 | Maternal dietary Se intake in the first 4-5 months of pregnancy was associated with higher birth weight and population-based birth weight z-scores (SD: 14.3 ug/day) of Se intake<br>Negative associations observed between birth weight and maternal whole blood Se, additionally non-significant but higher OR for SGA |
| Moody et al., 2020 | Community based cohort of n = 100 children 6-59 months | Venous blood Se (median, IQR) (ug/dL): 12.20 (10.69-15.02)<br>Stunted (median, IQR) (ug/dL): 11.53 (9.65-12.13)<br>Not stunted (median, IQR) (ug/dL): 13.05 (10.9-15.49) | $\beta$: 1.924 (p = 0.005) | Significant positive association between Se and HAZ score<br>Se showed a u-shaped toxicity dose-response curve, with negative effects at deficient and toxic levels |
| Guo et al., 2021 | Prospective birth cohort study n = 1931 pregnant women (28-36 gestational weeks) | Maternal serum Se: 136.9±47.9 ug/L | Birth weight (gms) (3.4±0.4 kg) $\beta$: 0.003 (-0.001, 0.006); -0.0002 (-0.0008, 0.00003)<br>Recumbent length (0.1 cm) (49.9±1.9 cm) $\beta$: 0.004 (-0.003, 0.012); 0.001 (-0.0002, 0.002)<br>Maternal Se levels: ≥ 103.7 ug/L (highest 3 quartiles) | Effect sizes were small and non-significant for the associations between maternal serum Se and birth weight, recumbent length, although associations were positive for Se concentrations in the highest 3 quartiles compared to the lowest Se concentration quartile |
| Zhang et al.,<br>2021 | China-Anhui Birth<br>Cohort Study – n =<br>3133 mother-infant<br>pairs | Maternal serum Se: 123 ug/L (2.0-440.6) | Birth weight (g): $\beta$: 0.58 (0.34,<br>0.82), $p < 0.0001$<br>Birth length (cm): $\beta$: 0.001<br>(0.000, 0.003), $p = 0.03$<br>Head circumference (cm): $\beta$:<br>0.0002 (0.000, 0.001), $p = 0.61$<br>Chest circumference (cm): $\beta$:<br>0.001 (0.000, 0.002), $p = 0.13$ | Odds of low birth weight were 3.92<br>(2.03, 7.57) times higher<br>OR for SGA were 2.77 (1.92, 4.02)<br>times higher among infants with<br>mothers who were selenium<br>deficient ($< 45$ ug/L)<br>ORs for LBW were 1.87 (1.02,<br>3.45) times higher and ORs for<br>SGA were 1.47 (1.07, 2.02) times<br>higher among infants with mother<br>who were selenium insufficient<br>(45-94.9 ug/L), all results were<br>statistically significant ($p < 0.001$) |

In a recent review of studies conducted across a variety of different geographies, results of the association between selenium and birth and growth outcomes have been mixed, although overall associations have suggested a positive and protective effect of this micronutrient (pooled correlation coefficient between VC blood Se and birth weight; r: 0.08, 95% CI: 0.01-0.16 and maternal blood Se and birth weight; r: 0.02 (−0.01-0.05) on birth weight).^6^ In our study, we found a strong negative association between whole blood Se in venous cord blood and birth weight (β: - 2.3, 95% CI: -3.7 to -0.98, p-value: 0.001); however, the association between SEPP1 in venous cord and birth weight ran in the opposite direction (β: 0.04, 95% CI: -0.01 to 0.1, p-value: 0.1). Our findings negate our hypothesis that SEPP1 may mediate the association between venous cord WBSe and weight related outcomes (birth weight, WAZ at birth and at 1-year of age).

The negative association between whole blood Se in venous cord blood and birth weight may be a result of high selenium exposure in utero. This can lead to oxidative stress, which in turn causes hypoxia and can impact fetal growth by reducing the distribution of oxygen in the placenta.^46-47^ It is important to note that selenium concentrations at levels classified as toxic were not detected in any of our specimens (> 400 μg/L).^30^ Non-selenoprotein bound Se concentrations are known to be higher in the placentas of women who experience intrauterine growth restriction (IUGR) and in mothers of preterm infants.^9^ These findings add further credence to our observations around the negative associations between WBSe and birth weight.^47^

The protective antioxidant effects of selenium bound to selenoproteins including SEPP1 may work to increase birth weight during gestation. This may explain the positive associations with birth weight as observed in our study, given the known antioxidant properties of selenoproteins, which can protect the fetus from free radicals in utero, modulate inflammation and protect biological membranes and DNA during early stages of embryo development.^6^ As expected, a plateauing effect was noted in our results, for the association between WBSe and functional selenoprotein, SEPP1 in venous cord blood at WBSe concentrations of approximately 160 μg/L; however no distinct plateauing was observed for associations with delivery biomarkers. Mixed findings about the effects of varied biomarkers of Se status on placental development and growth-related outcomes, postpartum, may be explained by differences in bioavailability of organic and inorganic forms of the micronutrient.^30^ This is important because absorption of these forms of Se, once in the bloodstream, can impact distribution, partitioning and potential toxicity at high concentrations.^30^

The primary mechanisms whereby selenium has been associated with adverse growth outcomes include its essential role in antioxidant defense and its role in mediating oxidative stress, which could impact perinatal morbidity and mortality.^9^ This in turn may impact risk of outcomes such as miscarriage, pre-eclampsia, gestational diabetes, and IUGR.^9^

Low selenium status, particularly in women, can impact the risk of PTB.^9^ Our results are in contrast to these findings however, where we found higher odds of PTB per unit increase in maternal delivery whole blood and serum Se and WBSe in venous cord blood.^9^ These findings were not significant after accounting for confounding variables. Associations have been noted between lower serum Se and PTB (OR: 2.0, 95% CI: 1.19-3.47).^48^ On the other hand, the risk of PTB was lower per unit increase in SEPP1 in maternal delivery and venous cord blood specimens, although our results did not approach statistical significance.

Mechanistically, selenium and PTB are linked because of the role this micronutrient plays in the body’s inflammatory response which can in turn cause ruptures to the fetal membrane.^48^ Serum selenium concentrations that surpass 80.5-86.9 μg/L can cause an attenuation in the odds of PTB.^48^ We noted similar findings in our study, where we saw an attenuation in ORs for the highest quintile of serum selenium concentrations in our sample (OR: 1.3, 95% CI: 0.7, 2.4) compared to the lowest, and a decreasing trend in ORs by increasing quintiles of serum Se. Women at increased risk of miscarriage may also have lower serum Se concentrations, particularly during the first trimester of pregnancy.^49^ Furthermore, cord blood Se concentrations are known to be significantly lower in premature infants when compared to neonates born at term.^50^

Similar to other birth-related outcomes, an increased risk for SGA has also been observed in relation to selenium, likely driven by mechanisms associated with oxidative stress.^51^ We found a per unit increase in concentrations of several selenium biomarkers to result in a small and non-statistically significant increase in the risk of SGA. It is likely that inflammation plays a role in our findings, given that median concentrations of CRP at delivery were 9 mg/L, which are higher than clinically defined thresholds for this acute phase protein (< 5 mg/L).^52^ We have therefore adjusted our models to account for potential confounding. We noted an attenuation in the odds of LBW across quintiles of maternal serum selenium at delivery, which suggest that higher concentrations of selenium are associated with lower odds of LBW (OR: 0.89, 95% CI: 0.6, 1.4), which is consistent with other studies.^41^

In our study, we found very small negative associations between WBSe in venous cord blood and LAZ across age time points, in unadjusted and adjusted linear models, although other selenium biomarkers were not associated with LAZ. We observed small effect sizes that were not significant for associations between serum selenium at delivery and LAZ. This was in line with the literature for maternal serum selenium.^19, 41^

Linear growth has been implicated as a potential outcome of interest in relation to the role of selenium in bone growth. Kashin-Beck syndrome (KBD) is a chronic osteochondropathy known to result from inherited genetic polymorphisms in selenium-related genes which impair collagen synthesis.^53-54^ The syndrome is endemic to regions with low soil selenium concentrations.^53^ A key feature of the condition is necrosis of chondrocytes in the growth plate of the bone and within the articular surface, which results in growth retardation and osteoarthritis.^55-56^ In this condition, selenium status may modulate disease susceptibility because of its essential antioxidant activities. Three major environmental hypotheses have been postulated as part of the etiology of the condition and include, soil selenium deficiency, contamination of cereals by mycotoxigenic fungi and high humic acid concentrations in the drinking water supply in endemic areas including China and Tibet.^57^ In a clinical study conducted in Lhasa, Tibet, where KBD is endemic and selenium deficiency is common, authors found serum Se concentrations to be below detectable levels (< 5 μg/L) in 38% of the study sample (n = 521 subjects). Geometric mean concentrations of serum selenium in this study sample for subjects who had KBD were as low as 10.3 (5.1-20.8) μg/L. These values are significantly below those observed in our study from Bangladesh, where we found mean concentrations of maternal serum Se to be 69 ± 13 μg/L at delivery. These findings suggest that severe selenium deficiency may not be a problem in this Bangladeshi population (83.4% of our sample was < 80 μg/L for serum Se at delivery and only 0.2% were selenium deficient [< 30 μg/L]).

It is important to note that effect estimates for other growth outcomes in our study were negligible in magnitude and therefore do not suggest that selenium biomarkers measured in our study play a major role in birth and growth outcomes in this cohort of mother-infant pairs. We did however find meaningful and substantial effect sizes for the associations between birth weight and biomarkers of selenium status. These associations should be investigated further given that different selenium biomarkers may act differently on mechanisms of weight gain at birth and in early infancy.

Evidence exists to suggest that selenium has a U-shaped dose-response toxicity curve, with negative health impacts experienced due to deficiency and excess as is evidenced in our dose-response analyses as well.^58-59^ Selenium deficiency may contribute to inflammation and pregnancy related complications, via pathways associated with oxidative stress and may also be associated with growth retardation.^60^ At high concentrations, selenium can become a pro-oxidant and cause cellular damage due to oxidative stress.^60^ In conclusion, we have found that selenium may play an important role in placental health and the growth and development of infants and young children in the peri-natal and postpartum period, up to two years of age. Future studies should examine in more detail, the role of selenium in linear growth over the infant and young child life course, particularly in relation to its important known functions in placental preservation and birth weight.

## Data Availability

All data produced in the present study are available upon reasonable request to the authors

## Contributions

RM and CK designed the study, RM conducted analysis and wrote the manuscript. DHH provided conceptual inputs to this study and critical review of the manuscript. TM and AAM oversaw the implementation of the MDIG trial in Dhaka, Bangladesh. All authors have read and approved of the final version of this manuscript.

## Acknowledgements

We wish to thank Dr. Daniel E. Roth, PI of the MDIG Trial for providing conceptual inputs to this study, review of methods and results and access to the MDIG data. We thank Huma Qamar, Ulaina Tariq and the SickKids team for their contributions to methods and review of findings and overall contributions to the MDIG study as well as Jithin Sam Varghese for consultation on analytical methods and interpretation. We wish to acknowledge the teams at ICDDR, B, and other MDIG collaborators in Bangladesh and internationally for all their contributions to the parent study. We also acknowledge the CDC for selenium biomarker analyses. Finally, we extend our gratitude to the Bangladeshi mothers and children who participated in this study.

---

**Supplementary Figure 1:**
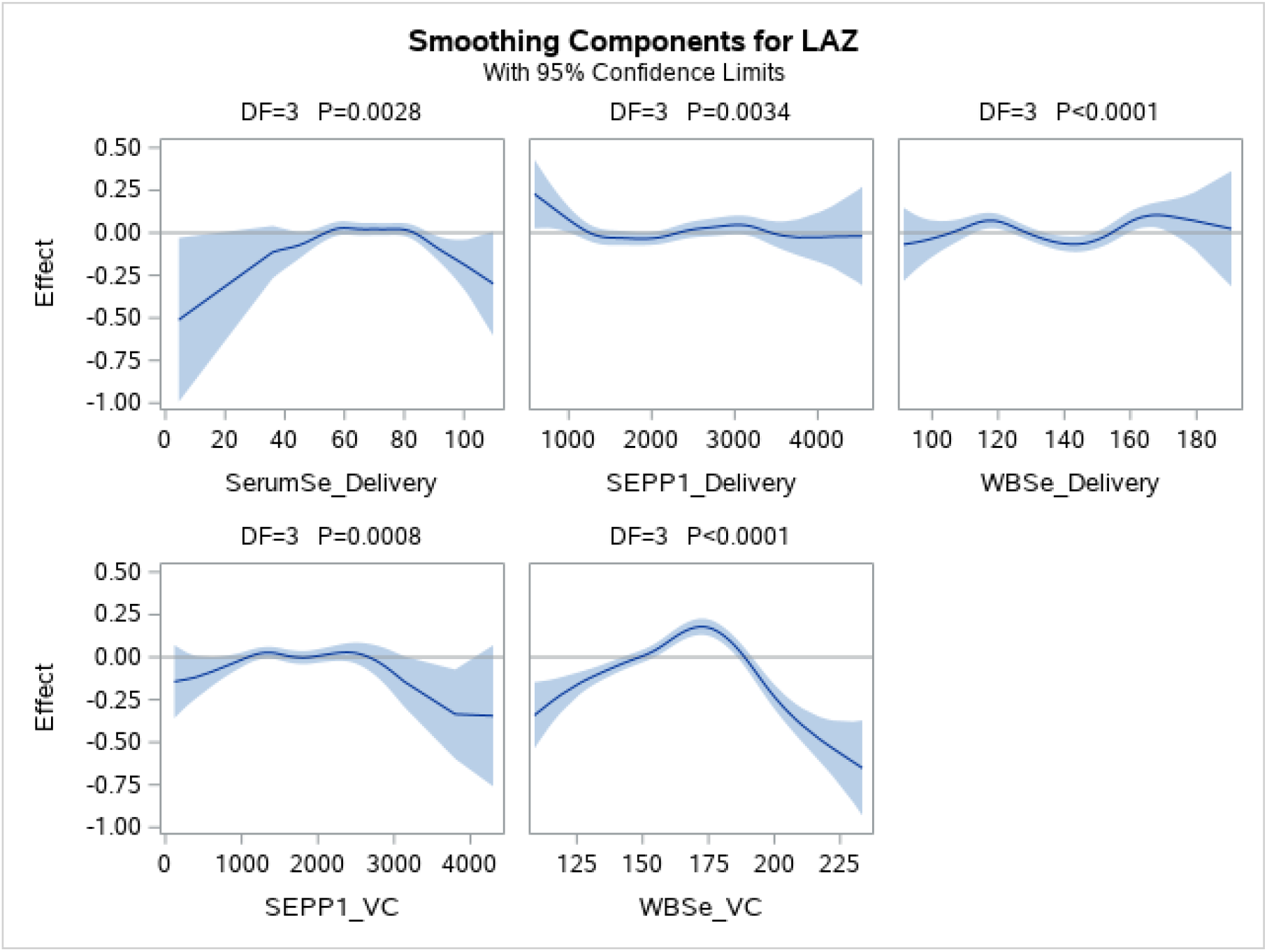
General additive models for associations between length-for-age z-scores and selenium biomarkers at delivery and in venous cord blood

**Supplementary Figure 2:**
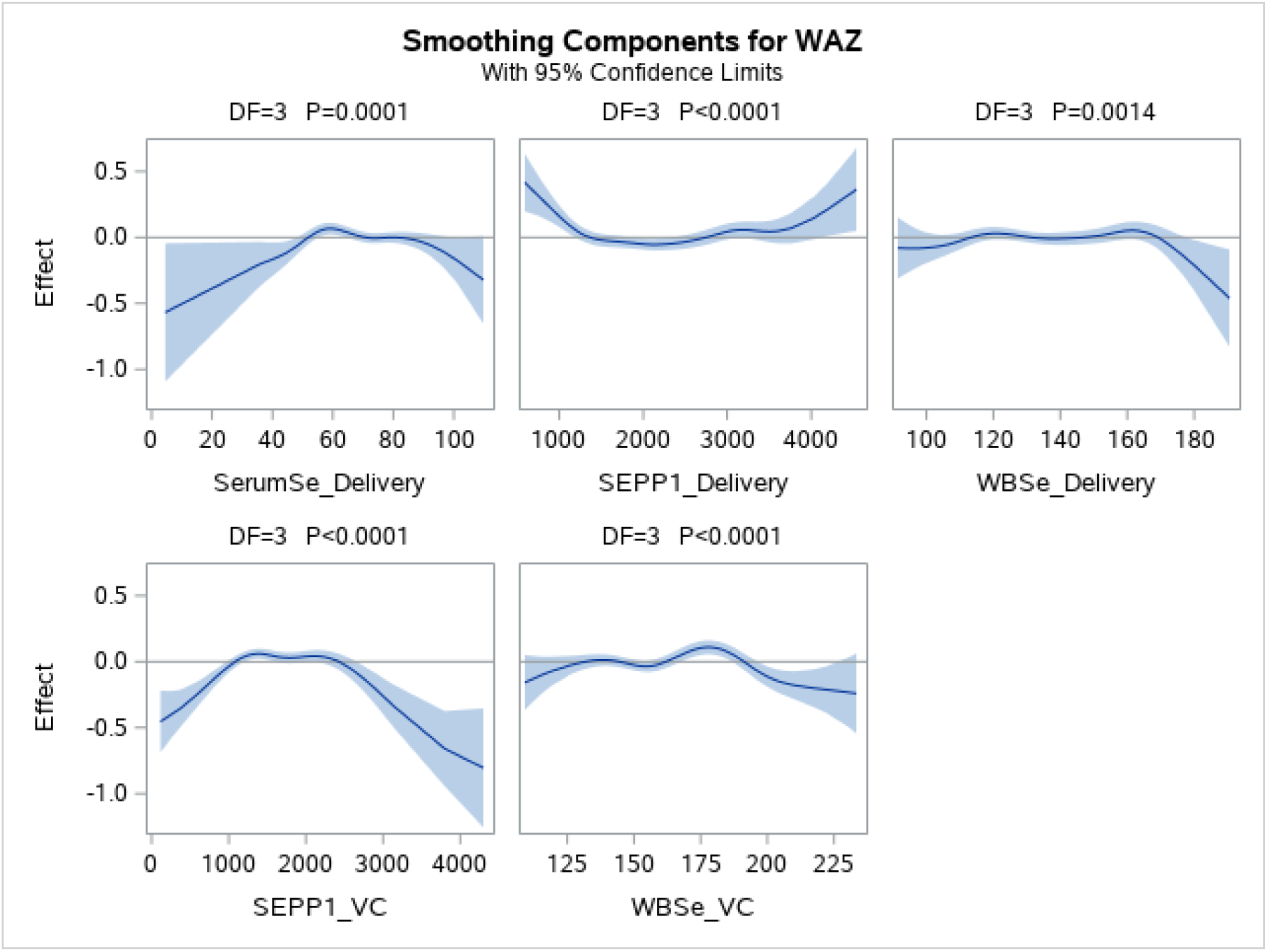
General additive models for associations between weight-for-age z-scores and selenium biomarkers at delivery and in venous cord blood

**Supplementary Figure 3:**
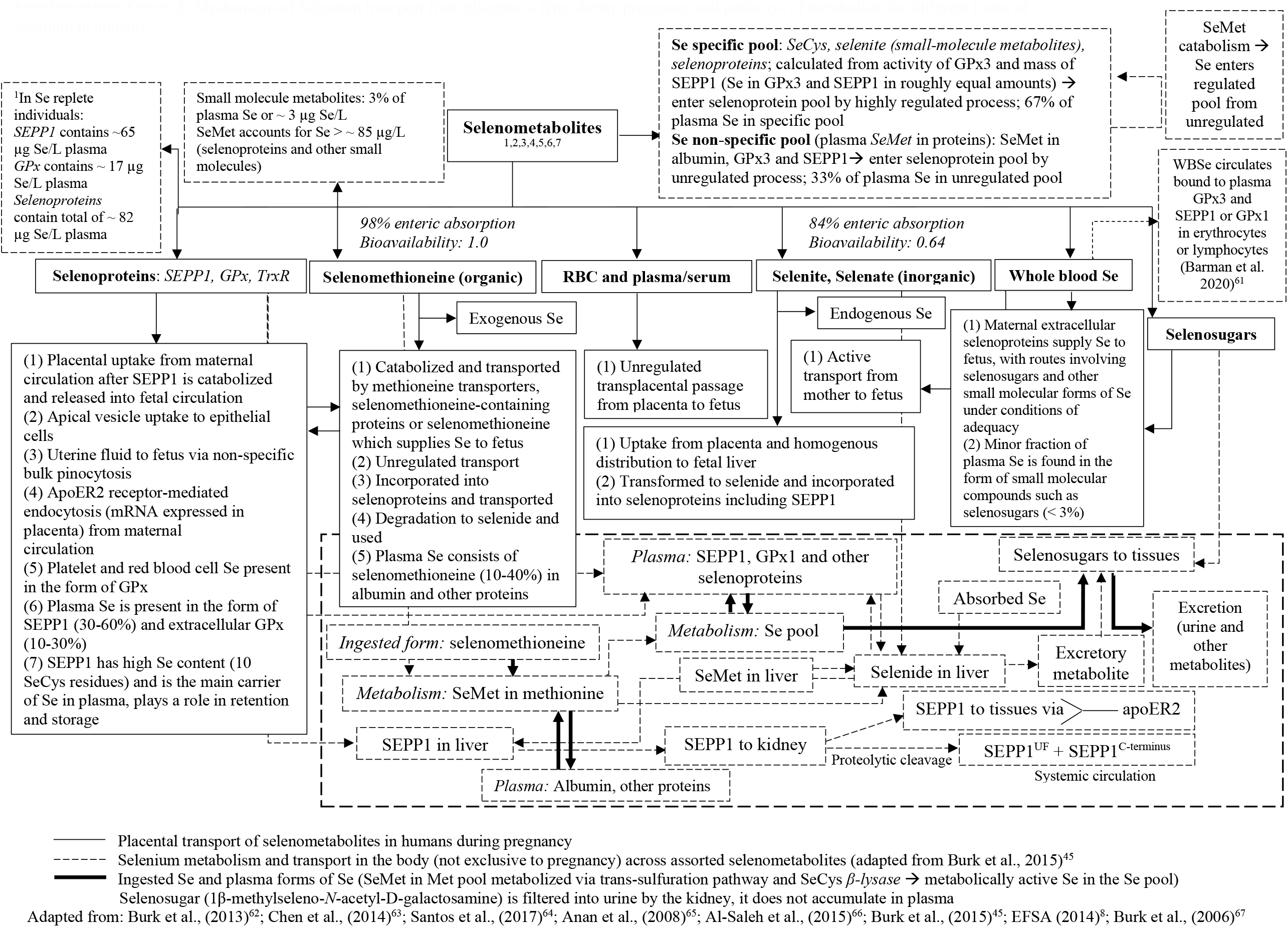
Mechanisms of Selenium transport from placenta to fetus during pregnancy and pathways of metabolism for different forms of selenium in humans

**Supplementary Table 1:** Selenoprotein concentrations in mid-gestation and delivery specimens in the growth study

| **Biomarkers** | **Maternal mid-gestation (17-24 weeks)** |  | **Maternal delivery** |  |
| --- | --- | --- | --- | --- |
|  | **GPx** | **TrxR** | **GPx** | **TrxR** |
| *N*<sup>1</sup> | *201* | *201* | *145* | *144* |
| **Selenium biomarker concentration (µg/L), median (IQR)** | 135.3 (32.3) | 4.8 (2.8) | 138.3 (36.8) | 7.3 (8.1) |
| Gestational age at birth (in weeks), median (IQR) | - | - | 38.9 (1.9) | 38.9 (1.9) |
| Birth weight (g), mean (SD) | - | - | 2647.5 (470.0) | 2653.8 (481.3) |

<sup>1</sup>Sample sizes exclude observations where measures of selenoprotein & birth weight are not available

**Supplementary Table 2:** Linear associations between selenoproteins – GPx and TrxR and birth weight

| Outcome | Se Biomarker | Timepoint | N | Unadjusted $\beta$<br>(95% CI) | p-value | N | Adjusted $\beta^2$<br>(95% CI) | p-value | Cotinine Sensitivity Analysis | | |
| --- | --- | --- | --- | --- | --- | --- | --- | --- | --- | --- | --- |
| | | | | | | | | | N | Adjusted $\beta^2$<br>(95% CI) | p-value |
| Birth weight<sup>1</sup> | GPx | Mid-gestation | 145 | 1.1 (-0.2, 2.3) | 0.09 | **140** | **1.3 (0.1, 2.5)** | **0.04** | 88 | 1.0 (-0.7, 2.6) | 0.2 |
|  |  | Delivery | 145 | -0.7 (-1.8, 0.4) | 0.2 | **141** | **-1.1 (-2.1, -0.07)** | **0.04** | 90 | -0.6 (-2.0, 0.8) | 0.4 |
|  | TrxR | Mid-gestation | 145 | 4.6 (-8.2, 17.5) | 0.5 | 140 | 10.6 (-1.6, 22.8) | 0.09 | 88 | 15.6 (-3.4, 34.6) | 0.1 |
|  |  | Delivery | 144 | -0.1 (-6.9, 6.7) | 0.9 | 140 | 0.09 (-7.0, 7.1) | 0.9 | 89 | 1.6 (-7.5, 10.6) | 0.7 |

<sup>1</sup>No scaling applied to GPx and TrxR biomarkers
<sup>2</sup>Models adjusted for infant sex, gestational age at birth, season of birth, maternal age, maternal education, maternal height, maternal BMI, gravidity, vitamin D treatment group, delivery CRP, asset index quintiles, daily protein intake (kg), smoking and tobacco use during pregnancy, cotinine included in sensitivity analysis

**Supplementary Table 3:**
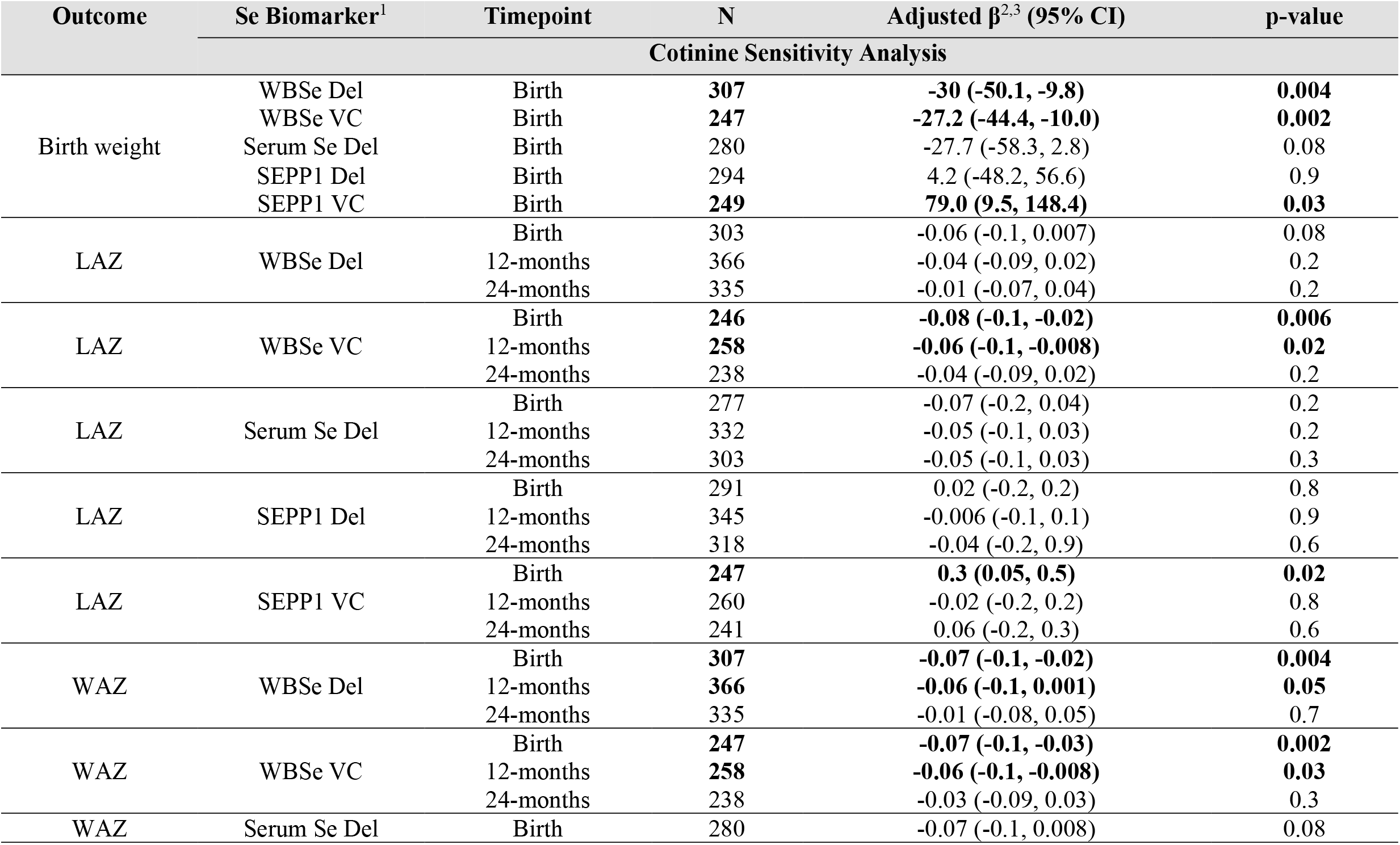
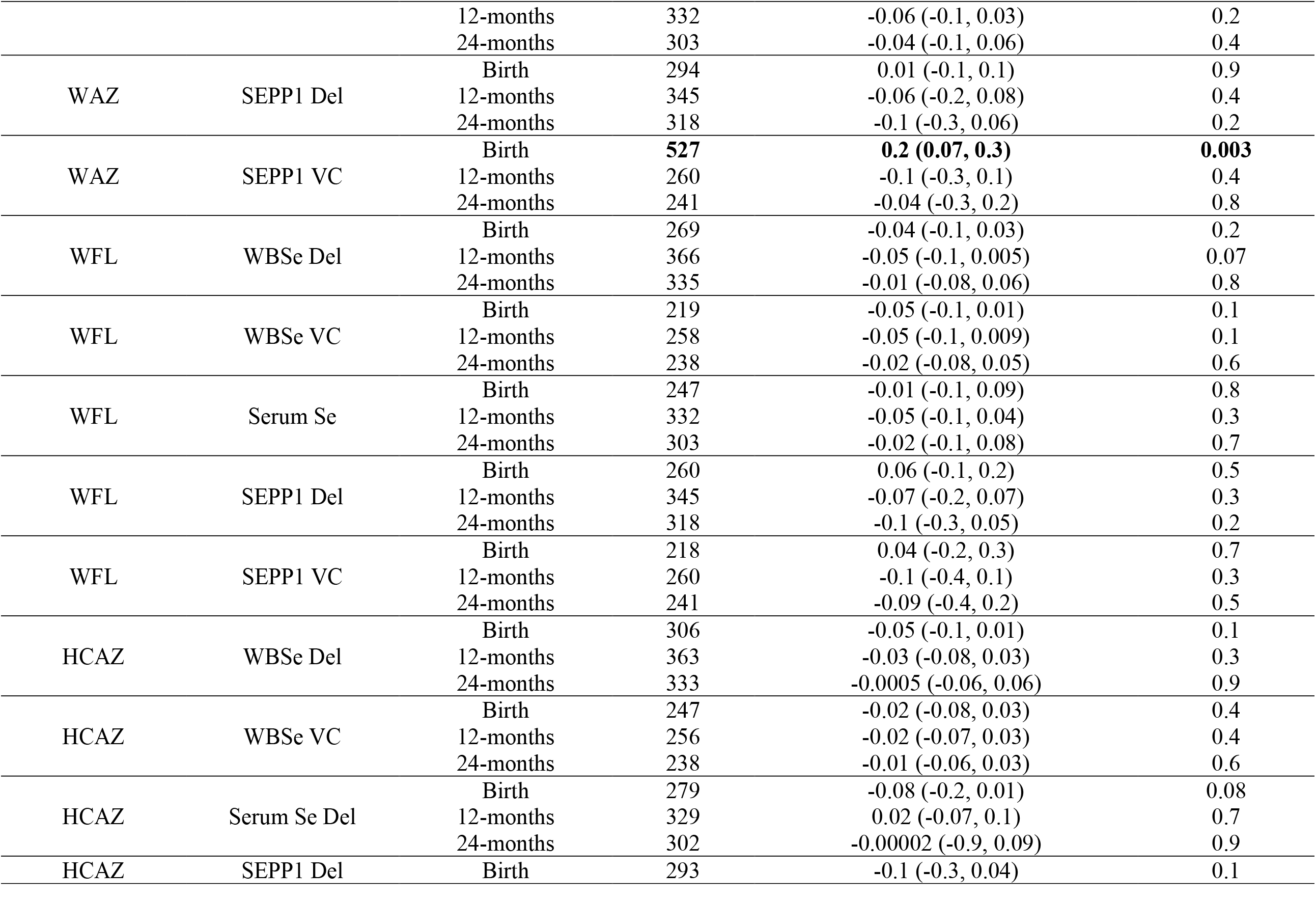
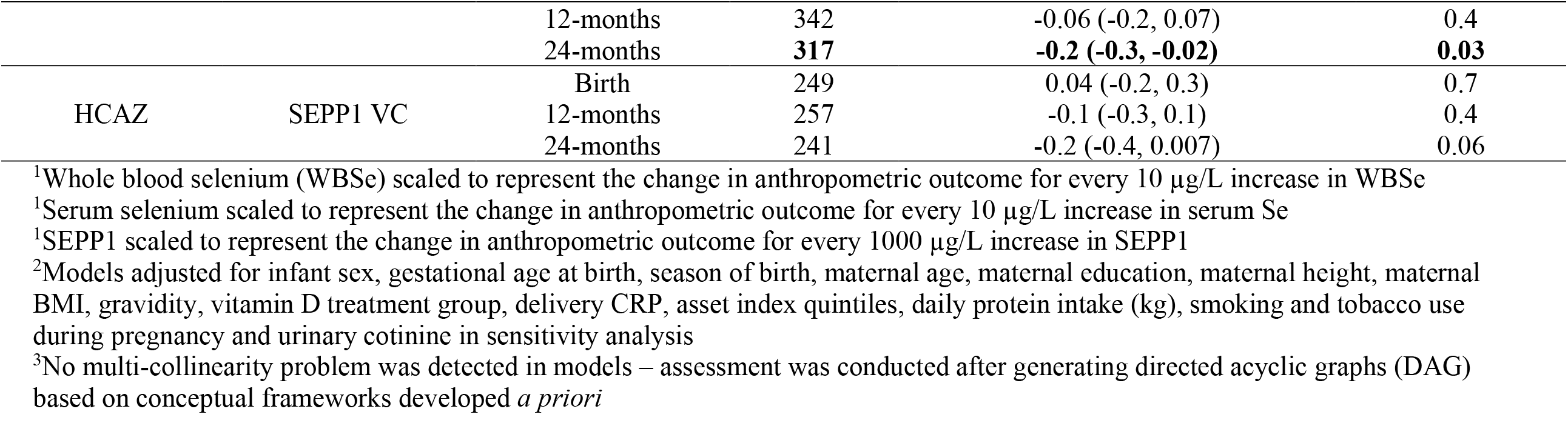
Linear regression sensitivity analyses for associations between maternal delivery and venous cord Selenium biomarkers and growth outcomes adjusting for urinary cotinine

**Supplementary Table 4:** Modified Poisson Regression sensitivity analyses for associations between maternal delivery and venous cord Selenium biomarker and birth outcomes adjusting for cotinine analysis

| Outcome | Se Biomarker | Timepoint | N | Adjusted Risk Ratio (95% CI) | p-value |
| --- | --- | --- | --- | --- | --- |
| Cotinine Sensitivity Analysis |  |  |  |  |  |
| Small-for-gestational age (SGA) | WBSe Del | Delivery | 307 | 1.06 (0.96, 1.2) | 0.3 |
|  | WBSe VC | Venous Cord | 247 | 1.10 (1.0, 1.2) | 0.02 |
|  | Serum Se Del | Delivery | 280 | 1.11 (0.96, 1.3) | 0.2 |
|  | SEPP1 Del | Delivery | 294 | 0.98 (0.75, 1.3) | 0.9 |
|  | SEPP1 VC | Venous Cord | 249 | 0.8 (0.6, 1.2) | 0.3 |
| Low birth weight (LBW) | WBSe Del | Delivery | 307 | 1.15 (1.0, 1.3) | 0.03 |
|  | WBSe VC | Venous Cord | 247 | 1.12 (1.0, 1.2) | 0.06 |
|  | Serum Se Del | Delivery | 280 | 1.18 (0.97, 1.4) | 0.1 |
|  | SEPP1 Del | Delivery | 294 | 1.12 (0.78, 1.6) | 0.5 |
|  | SEPP1 VC | Venous Cord | 249 | 0.9 (0.6, 1.5) | 0.8 |
| Preterm birth (PTB) | WBSe Del | Delivery | 308 | 1.42 (0.9, 2.1) | 0.08 |
|  | WBSe VC | Venous Cord | 248 | 0.99 (0.7, 1.5) | 0.9 |
|  | Serum Se Del | Delivery | 281 | 1.65 (0.96, 2.8) | 0.07 |
|  | SEPP1 Del | Delivery | 295 | 1.64 (0.63, 4.3) | 0.3 |
|  | SEPP1 VC | Venous Cord | 250 | 1.4 (0.3, 7.3) | 0.7 |

<sup>1</sup>Whole blood selenium (WBSe) scaled to represent the change in anthropometric outcome for every 10 µg/L increase in WBSe
<sup>1</sup>Serum selenium scaled to represent the change in anthropometric outcome for every 10 µg/L increase in serum Se
<sup>1</sup>SEPP1 scaled to represent the change in anthropometric outcome for every 1000 µg/L increase in SEPP1
<sup>2</sup>Models adjusted for infant sex, gestational age at birth, season of birth, maternal age, maternal education, maternal height, maternal BMI, gravidity, vitamin D treatment group, delivery CRP, asset index quintiles, daily protein intake (kg), smoking and tobacco use during pregnancy and urinary cotinine in sensitivity analysis
<sup>3</sup>No multi-collinearity problem was detected in models – assessment was conducted after generating directed acyclic graphs (DAG) based on conceptual frameworks developed *a priori*

